# Vaccine effectiveness against mild and severe disease in pregnant mothers and their infants in England

**DOI:** 10.1101/2023.06.07.23290978

**Authors:** Freja C M Kirsebom, Nick Andrews, Anna A Mensah, Julia Stowe, Shamez N Ladhani, Mary Ramsay, Jamie Lopez Bernal, Helen Campbell

## Abstract

**Background:** Pregnant women are at increased risk of adverse outcomes following SARS-CoV-2 infection, including venous thromboembolism, admission to critical care and death. Their infants may also suffer from pre-term birth, stillbirth and severe disease. Vaccination may protect both mothers and their infants against severe COVID-19 disease.

**Methods:** We used a test-negative, case-control study design to estimate vaccine effectiveness against symptomatic disease and hospitalisation with the Delta and Omicron variants in pregnant women who gave birth in 2021 in England. We also estimated the protection conferred by prior infection and maternal vaccination against symptomatic disease and hospitalisation with the Delta and Omicron variants in their infants.

**Findings:** Vaccine effectiveness against symptomatic disease (Delta and Omicron) and against hospitalisation (Delta only) was high and similar to that observed in the general population. Maternal vaccination during and post-pregnancy as well as previous infection also provided sustained protection from symptomatic disease and hospitalisation following Delta and Omicron infection in infants up to 8 months of age, with the highest protection being observed when maternal vaccination occurred during later pregnancy. Unlike non-pregnant women, a booster dose provided sustained protection with no evidence of waning up to 15 weeks after vaccination.

**Interpretation:** Maternal vaccination prevents mild and severe disease in pregnant women and their infants up to 6-8 months after birth. Our findings support the promotion of both primary and booster vaccination for pregnant women, irrespective of prior infection status, to protect themselves and their infants.

**Funding:** None.

**Research in Context:** *Evidence before this study:* Pregnant women were included in the UK’s priority risk groups for COVID-19 vaccination from 2 December 2021 when they were encouraged to complete vaccination with an mRNA booster vaccine of either Pfizer BioNTech or Moderna. We searched PubMed using the terms ‘pregnancy’, ‘COVID-19’, ‘vaccine’ and ‘vaccine effectiveness’, with no date restrictions on 1 March 2023, and used the snowball process to identify additional relevant publications. We also scoped preprint databases for relevant COVID-19 vaccine effectiveness studies undertaken after the emergence of the more immune-evasive Omicron variant from December 2021. Studies have shown moderate COVID-19 vaccine effectiveness after a second dose in pregnant women against symptomatic Omicron disease with evidence that booster doses of mRNA vaccines confer higher protection against serious Omicron disease, comparable with population-based immunity. In addition to evidence of transplacental transfer of maternal antibody, real-life evidence from test-negative case-control studies have demonstrated protection in infants following maternal vaccination which is highest after vaccination in the third trimester and wanes with increasing infant age.

*Added value of this study:* Ours is the largest study of the effectiveness of maternal COVID-19 vaccines against both maternal and infant disease, in addition to the protection conferred by past infection in the mother to the infant. In pregnant women, vaccine effectiveness against symptomatic Delta and Omicron infection, and against hospitalisation with Delta, remained high after vaccination with limited waning observed at the longest time points investigated post vaccination. Both prior infection and maternal vaccination protected infants after birth against symptomatic disease and hospitalisation with Delta and Omicron. Vaccine effectiveness was highest when maternal vaccination occurred in the later stages of pregnancy.

*Implications of all the available evidence:* These findings support the benefits of maternal vaccination in preventing disease in the mother and in her infant in the first months of life, regardless of prior infection status in the mother. Policy decisions need to balance the suggestion of higher protection after vaccination later in pregnancy with the need to ensure adequate opportunities for vaccination before women reach the pregnancy stage when they are at greatest risk from COVID-19 disease and to optimise the infant benefit even in pre-term births.

## Introduction

Pregnant women are at increased risk of adverse outcomes following SARS-CoV-2 infection, including venous thromboembolism and admission to critical care, compared to their non-infected pregnant peers, as well as increased risk of pre-term birth (spontaneous and iatrogenic) compared to pregnant women without COVID-19 (1-5). Higher rates of respiratory support, ICU admission and death were reported in US women giving birth with a COVID-19 diagnosis recorded in the same admission compared to other pregnant women (6). Increased risk of stillbirth and neonatal mortality has also been reported in babies born to women with SARS-CoV-2 infection in later pregnancy, as well as increased odds of neonatal admission to intensive care (3). COVID-19 was ranked as seventh leading cause of death in infants under 1 year of age in the US accounting for 0.7% of all deaths in this age group (7). In contrast, studies have consistently found no association between COVID-19 vaccination in pregnancy and adverse outcomes in pregnant women, the pregnancy, or the neonate (1, 2, 8-12).

COVID-19 vaccine in pregnancy can directly reduce the risk of serious disease in pregnant women. Vaccination can thereby protect babies *in utero* against the consequences of serious disease in the mother, such as preterm birth and stillbirth (10). There is also growing evidence that booster doses of mRNA vaccines offer increased protection against severe Omicron disease requiring hospitalisation or emergency department/urgent care support in pregnant women when compared to 2-doses or to unvaccinated women (13, 14). Recent findings from a multinational study in the Omicron era found 2-dose vaccine effectiveness in women diagnosed with COVID-19 [VE] of 74% (95% confidence interval [CI], 48-87%) against severe complications, rising to 91% (95%CI, 65-98%) after a booster dose (15).

In addition, transplacental transfer of maternal antibodies can potentially provide direct passive protection in the infant from birth in their first months of life. There is evidence of maternal vaccine-elicited antibodies against SARS-CoV-2 in infant cord blood (2, 16), and real-world evidence from test-negative case-control studies, which found 2 and 3-dose maternal vaccination also protected infants against infection and hospitalisation (17-20). Protecting the mother through vaccination may also reduce her risk of SARS-CoV-2 infection and consequently reduce her risk of infecting her infant after birth.

As part of the UK COVID-19 vaccination programme, from December 2020, adults in England were offered a primary course of two doses of either Oxford/AstraZeneca (ChAdOx1-S), Pfizer BioNTech (Original Comirnaty®) or Moderna (Spikevax®) and an mRNA booster vaccine of either Pfizer BioNTech (Original Comirnaty®) or Moderna (Spikevax®). From 16 April 2021, the Joint Committee on Vaccination and Immunisation (JCVI) advised that pregnant women be offered COVID-19 vaccines at the same time as people of the same age or risk group (21). Under ongoing programme review, the JCVI met on 2 December 2021 and considered further data on severity of SARS-CoV-2 infection in pregnant women together with data on vaccine safety, which led to inclusion of pregnant women in the UK’s priority COVID-19 vaccine list and encouragement to complete vaccination (22).

Here, we estimate VE against symptomatic disease and hospitalisation with the Delta and Omicron variants in pregnant women who gave birth in 2021 in England using a test-negative case-control study design. Furthermore, we estimate the protection conferred by prior infection and maternal vaccination against symptomatic disease and hospitalisation with the Delta and Omicron variants in infants.

## Methods

### Study design

A test-negative case-control study design was used to estimate the effectiveness of the COVID-19 vaccines in preventing disease in pregnant women and to estimate the effectiveness of maternal vaccination and maternal prior infection in preventing disease in infants. Pregnant women who test positive for SARS-CoV-2 are defined as cases and those testing negative as controls, and the exposure of interest was either vaccination or prior infection.

SARS-CoV-2 PCR testing in England is undertaken by hospital and public health laboratories (Pillar 1), as well as by community testing (Pillar 2) (Supplementary Table 1). To estimate VE against symptomatic disease in pregnant women, the odds of vaccination in symptomatic Pillar 2 PCR-positive cases were compared to the odds of vaccination in symptomatic Pillar 2 PCR negative controls, as previously described (23-25).

To estimate the protective effect of maternal vaccination and prior infection against symptomatic disease in infants, Pillar 1 PCR tests in associated with a hospital admission with a respiratory infection in the primary diagnosis field were included as well as symptomatic Pillar 2 PCR tests, and the exposures of interest were maternal vaccination status and maternal past infection status.

To estimate VE against hospitalisation in pregnant women and infants, the odds of vaccination (or maternal vaccination) in hospitalised cases (SARS-CoV-2 Pillar 1 tests positive) which could be linked to a respiratory hospital admission with a minimum of a two-day in-patient stay) were compared to the odds of vaccination (or maternal vaccination) in negative controls, as previously described (24, 26, 27).

Analyses were stratified by Delta or Omicron variant.

Full details of data sources available in the Supplementary Appendix.

### Testing Data

Women who gave birth in 2021 and infants born in 2021 were linked to the Pillar 1 and 2 testing data by NHS number, with 58% of women and 40% of infants found to have been tested. For VE analysis of pregnant women, tests were included where the date of test was within 90 days prior to the pregnancy start date or up to 90 days after the delivery date.

### National Immunisation Management System (NIMS)

Testing data were linked to NIMS using combinations of the unique individual NHS number, date of birth, surname, first name, and postcode using deterministic linkage (Supplementary Table 1). NIMS was accessed for dates of vaccination and manufacturer, as well as demographic information and clinical risk status.

When assessing effectiveness in infants, maternal vaccination status was defined according to doses received by the mother more than 14 days before birth and doses received after this date but more than 14 days before the onset date in the infant. The trimester of the last dose a vaccine was received in pregnancy was defined as pre-pregnancy, trimester 1 (week 0(+0) to 11 (+6)), trimester 2 (week 12(+0) to 26 (+6), trimester 3 (week 27 (+0) to 15 days before birth). So, for example, an infant with onset aged 15 weeks with a mother vaccinated with a dose in trimester 2, a dose in trimester 3, and a dose 10 weeks after the birth would be coded as having had a second dose in trimester 3 and a dose post birth (short hand for figures: Tri3, dose2 +). Similarly, a mother given a first dose in trimester 2 and two doses after birth (and 14 days before onset) would be coded in figures as Tri2, dose 1 ++ where ++ means 2 doses after birth. When assessing effectiveness in the mother vaccination status was defined by number of doses at onset or sample date and the interval in weeks since the previous dose (0-1,2-4,5-9,10-14,15-19,20-24,25+) irrespective of whether the doses where actually given in pregnancy. Analysis was irrespective of manufacturer.

### Covariates and adjustment

Likely confounding variables were week of symptom onset, age (of infant and mother), risk group status of the women, region, IMD quintile, ethnicity and past positivity in the mother. In the infant analysis, additional confounding variables were pillar of testing (and its interaction with age), prematurity and sex. IMD quintile, region and ethnicity were assessed for confounding effects of 5% or more and only included if they changed effectiveness by at least this amount for at least one of the variants.

### Statistical method

Multivariable logistic regression was used with the test result as the outcome, vaccination status and past infection in the mother as the primary variables of interest and with confounder adjustment as described. VE or past infection effectiveness was calculated as 1-odds ratio and given as a percentage. Estimates are not shown where the number of controls was <30 or where the 95% CI lower bound was <-30% AND the top bound was >70%. The main comparator group was unvaccinated women or mothers never vaccinated (including after birth).

The primary analysis of VE during pregnancy included tests from the 3 months before the pregnancy start date up to the 3 months after giving birth. Sensitivity analyses were performed to assess VE just within trimester 1 to 3 of the pregnancy for the Delta period. Analyses were also conducted to estimate VE against symptomatic disease in all women aged 20 to 44 years tested in Pillar 2 during the Omicron period who were not pregnant.

In infants, the primary analysis included all infants 0 to 5 months of age. Stratification was by trimester of vaccination and also by infants age at onset 0 to 2 months and 3 to 5 months. For Omicron, it was also possible to do a stratification for infants aged 6 to 8 months.

## Results

### VE against symptomatic disease in pregnant women

Between 26 April 2021 and 9 January 2022 (the Delta variant study period) there were 35,210 negative and 16,693 positive eligible tests from symptomatic women aged 16-55 years who had given birth in 2021. Between 29 November 2021 and 31 March 2022 (the Omicron variant study period), there were 5,974 negative and 4,715 positive eligible tests. A description of eligible tests is included in Supplementary Table 3.

Following vaccination with a single dose of either ChAdOx1-S, BNT162b2 or mRNA1273, VE against symptomatic disease during pregnancy with the Delta variant was 50-60% and 30-40% with the Omicron variant, within 20 weeks of the dose being given (Table 1). After a second dose, protection against Delta peaked at nearly 90% then waned to about 60% after 25 weeks, whereas against omicron it peaked at 60% and waned to just below 30% over the same period. Booster vaccination with a third dose of either BNT162b2 or mRNA1273 increased VE against symptomatic disease with Delta to over 90% (based on small numbers) and Omicron to about 70% in the relatively short-term (most data <15 weeks) (Table 1, Figure 1.)

**Table 1.**
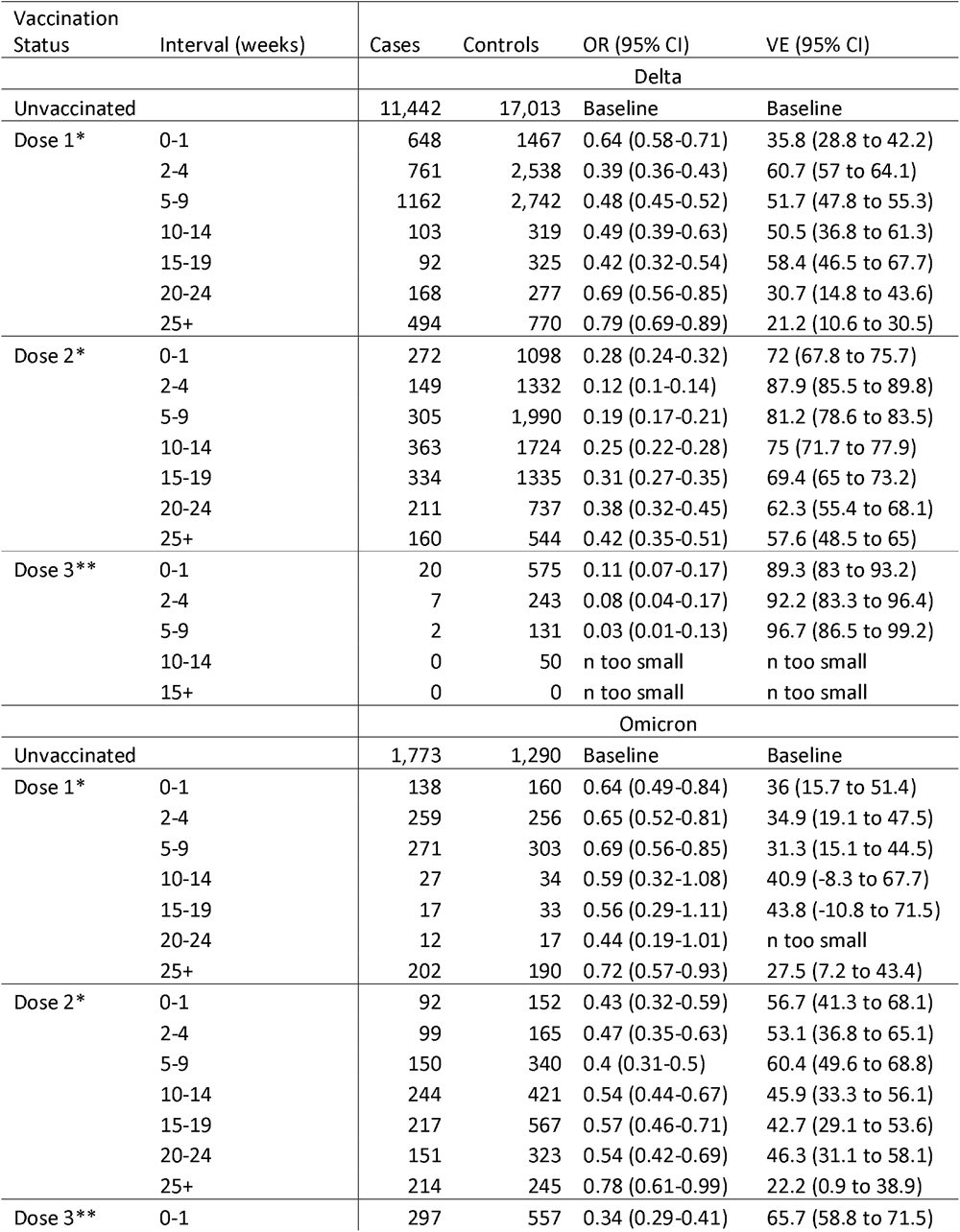
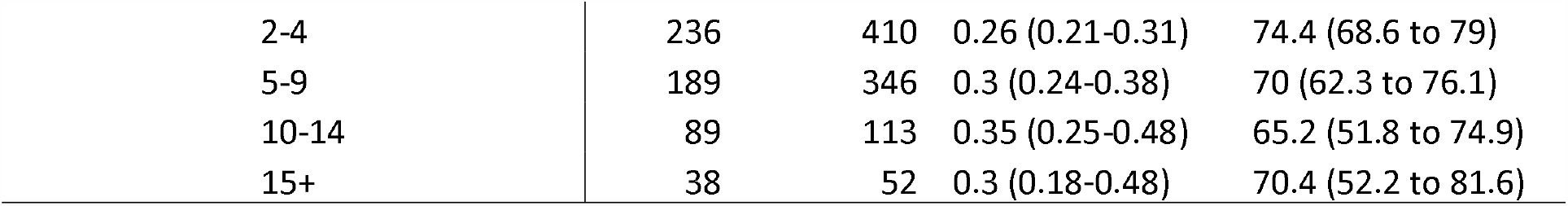
Vaccine effectiveness against symptomatic disease with Delta and Omicron in pregnant women within 3 months of their pregnancy start up to 3 months post-delivery. *Primary course manufacturer ChAdOx1-S, BNT162b2 or mRNA1273. **Booster manufacturer BNT162b2 or mRNA1273.

**Figure 1.**
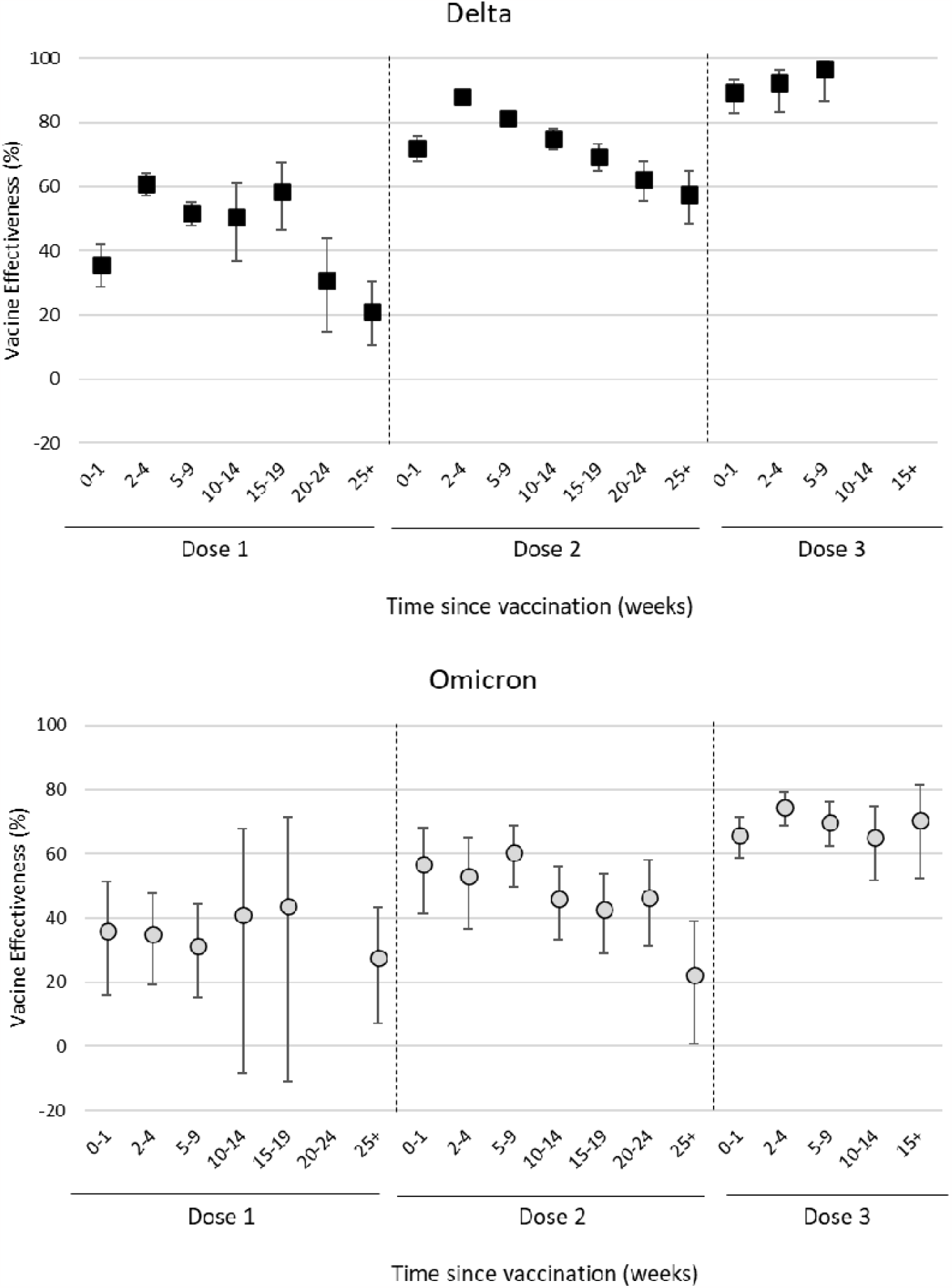
Vaccine effectiveness against symptomatic disease with Delta and Omicron in pregnant women within 3 months of their pregnancy start up to 3 months post-delivery, in England.

Sensitivity analyses including only the tests between the pregnancy start date and birth date (trimester 1 to 3) for the Delta period gave similar results (Supplementary Table 4). The analysis in non-pregnant women aged 20-44 years showed VE against symptomatic disease peaked at a similar level but waned more rapidly than pregnant women (Supplementary Table 5).

### VE against hospitalisation in pregnant women

In the Delta variant study period, there were 86 negative and 1,249 positive eligible tests from pregnant women tested in hospital settings (Pillar 1) which could be linked to a respiratory-coded hospital admission with a length of stay of at least two days (26) (Supplementary Table 6). There were insufficient data to estimate VE against hospitalisation with Omicron (11 eligible tests linked to a respiratory-coded hospital admission). VE against hospitalisation with Delta was >90% in the 5-19 weeks period after a first or second vaccine dose (Supplementary Table 7). There were insufficient data to estimate VE of a booster dose in the Delta or Omicron periods.

### Effectiveness of maternal vaccination and prior infection against symptomatic disease in infants with Delta and Omicron

Between 02 May 2021 and 09 January 2022 (the Delta variant study period) there was a total of 23,053 negative and 2,924 positive eligible tests from infants born in 2021, while between the 29 November 2021 and 31 March 2022 (the Omicron variant study period), there was a total of 13,908 negative and 5,669 positive tests. A description of infants with eligible tests is included in Supplementary Table 8.

Clear protective effects of maternal vaccination against symptomatic disease for both Delta and Omicron variants in infants aged 0 to 5 months were seen (Supplementary Table 9, Figure 2 & 3). The general pattern shows that doses given later in pregnancy gave higher protection with increasing protection after the 2^nd^ and 3^rd^ dose and that additional doses given to the mother after birth also added to the protection. VE was also higher against Delta than Omicron. The highest estimate against Delta was seen when the second dose was given in trimester 3 with VE at 86%. Numbers were too small to assess a third dose in trimester 3 for Delta, but for Omicron this gave a peak VE at 84%.

**Figure 2.**
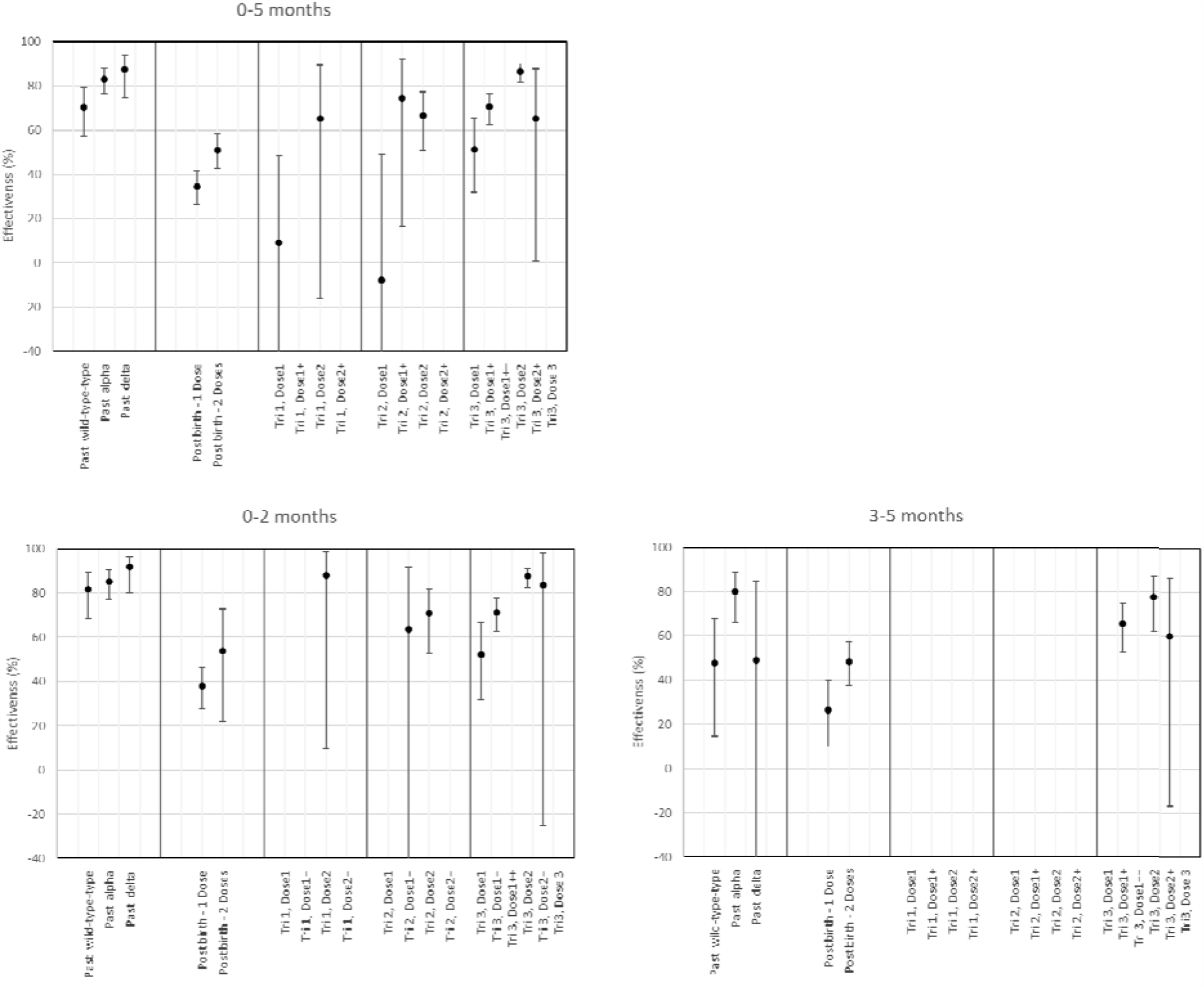
Protection of prior infection in the mother and maternal vaccination against symptomatic disease with Delta in infants in England.

**Figure 3.**
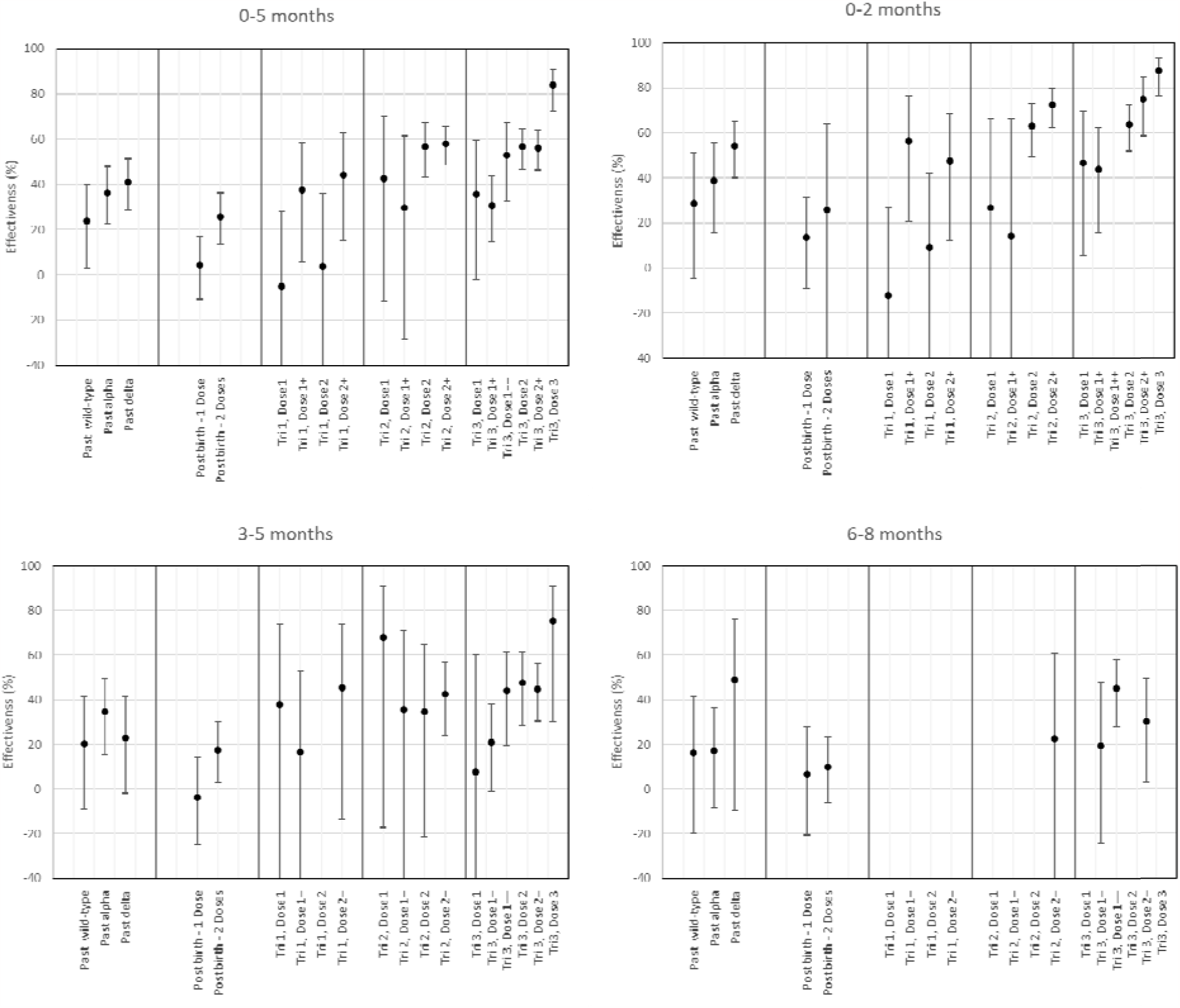
Protection of prior infection in the mother and maternal vaccination against symptomatic disease with Omicron in infants in England.

When stratified by infant age there was no significant reduction in the protection conferred by maternal vaccination against symptomatic disease with Delta or Omicron in infants aged 0 to 2 months as compared to infants aged 3 to 5 months. Longer term follow-up was unavailable for infants during the Delta period but maternal vaccination also had sustained protection against Omicron in infants aged 6 to 8 months, there was some suggestion of waning by this age, though confidence intervals overlapped (Supplementary Table 9, Figure 2 & 3). A protective effect in the infant was also observed where the mother was not vaccinated during pregnancy but was vaccinated after birth; the effectiveness of two doses post-delivery was around 50% and 25% against symptomatic disease with Delta and Omicron, respectively (Supplementary Table 9, Figure 2 & 3).

Prior COVID-19 infection in the mother also had a protective effect for the infant against developing symptomatic disease. Past maternal infection with wild-type, Alpha or Delta was around 70%, 83% and 88% protective, respectively, against infants aged 0 to 5 months developing symptomatic disease with Delta (Supplementary Table 10). Maternal infection had a reduced protective effect against symptomatic disease with Omicron than it did against Delta. The protective effect of past infection with wild-type, Alpha and Delta was around 24%, 37% and 41%, respectively, against Omicron in infants aged 0 to 5 months (Supplementary Table 10).

### Effectiveness of maternal vaccination and prior infection against infant hospitalisation with Delta and Omicron

Between the 2 May 2021 and 9 January 2022 (the Delta variant study period for infants, only tests from infants born after 2 May onwards were included as there was minimal maternal vaccination prior to this) there were 4,588 negative and 436 positive eligible tests in infants born in 2021, who were tested in hospital settings and linked to a respiratory-coded hospital admission with a length of stay of at least two days (26). Between 29 November 2021 and 26 June 2022 (the Omicron variant study period), there were 1,413 negative and 457 positive eligible tests (Supplementary Table 11).

Maternal vaccination during pregnancy and prior infection (during or prior to pregnancy) both protected infants from hospitalisation with Delta and Omicron, with higher VE against hospitalisation than against symptomatic disease (Supplementary Table 12 & 13, Figure 4). Protection conferred by maternal vaccination during the second trimester was around 70-90%, depending on the number of vaccine doses, for both Delta and Omicron. The highest protection in infants occurred when mothers were vaccinated during the third trimester; maternal vaccination with a second dose in the third trimester was approximately 95% and 78% protective against hospitalisation with Delta and Omicron, respectively. There were insufficient data to estimate VE of maternal boosters against severe disease in infants with Delta, but a maternal booster dose in the third trimester provided around 90% protection against hospitalisation with Omicron in infants aged 0-6 months (Supplementary Table 12 and Figure 4).

**Figure 4.**
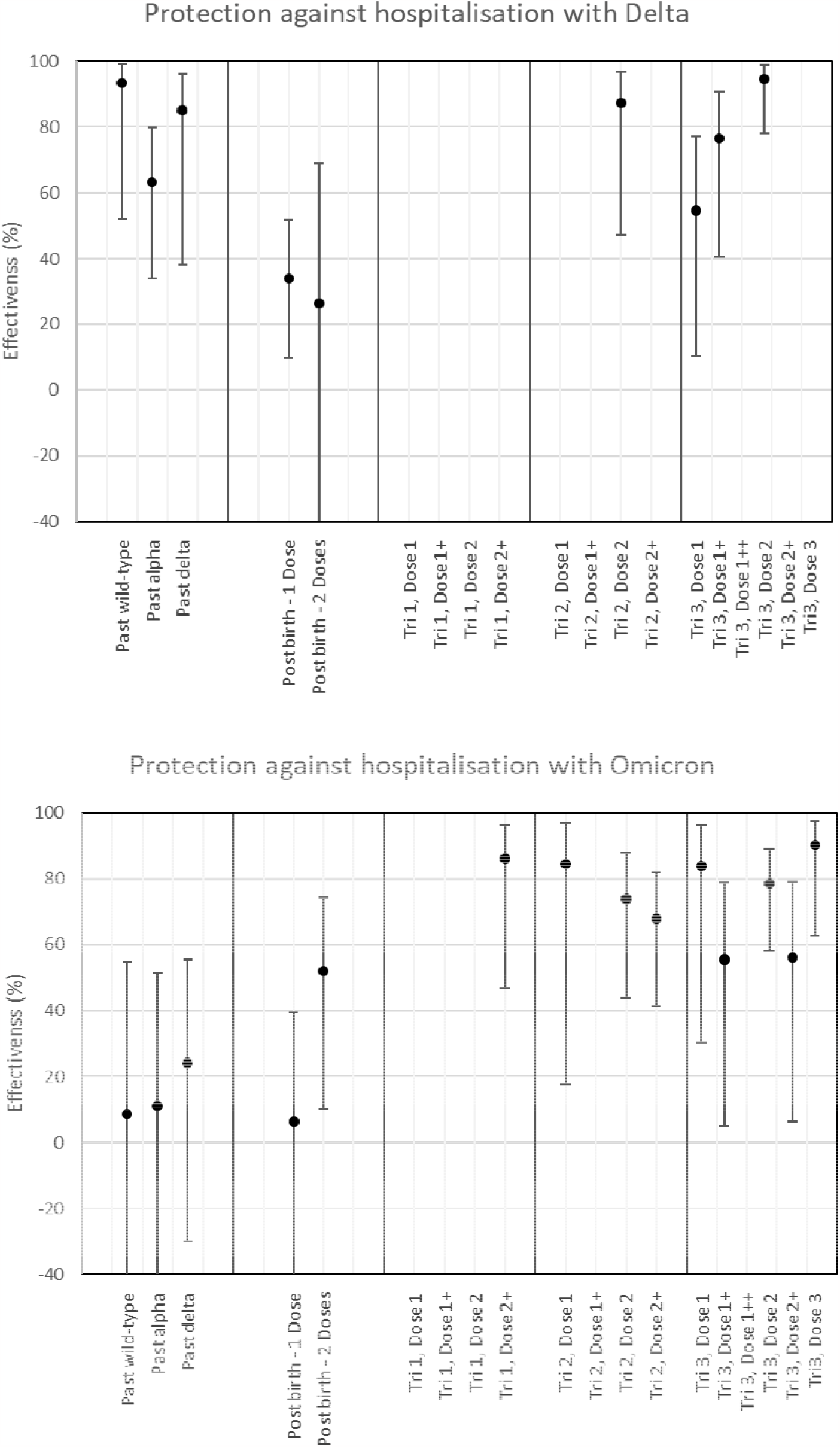
Maternal vaccine effectiveness against hospitalisation with Delta and Omicron in infants in England.

Prior maternal infection with any variant also protected infants from severe disease with Delta (Supplementary Table 13). The protective effect of previous maternal infection with wild-type, Alpha or Delta against infant hospitalisation with a Delta variant infection was 93.4% (95%CI, 52.0-99.1%), 63.3% (95%CI, 33.9-79.7%) and 85.1% (95%CI, 38.0-96.4%), respectively. The protection conferred by prior maternal infection against infant hospitalisation with Omicron was less evident with confidence intervals crossing 0%; the protective effect of previous wild-type, Alpha and Delta maternal infection against infant infection with Omicron was 8.4% (95% C.I.; -85.5-54.8%), 11.2% (95% C.I.; -61.9-51.3%) and 24.0% (95% C.I.; -30.1-55.6%) (Supplementary Table 13).

## Discussion

We find encouraging evidence that both prior infection in the mother and maternal vaccination (during and post-pregnancy) provides sustained protection against symptomatic disease and hospitalisation in the infant, with the highest protection being observed when maternal vaccination occurs during the later stages of the pregnancy. Protection against symptomatic COVID-19 and severe disease from prior infection or vaccination was significantly higher with the delta variant compared to the omicron variant.

Our VE estimates after the first and second dose against mild and severe disease with Delta variant in pregnant women was similar to earlier TNCC VE studies in the general population (15, 23, 28). Since the emergence of the more immune-evasive Omicron variant in late December 2021, we had limited data on VE after a third (booster) dose against Delta. VE against hospitalisation with Delta was very high, as we and others have observed previously (13, 26, 28). Sensitivity analyses demonstrated that including the period 3 months pre-pregnancy and 3 months post-birth yielded similar VE estimates compared to including the pregnancy period itself. Therefore, tests from these periods were used to estimate VE in pregnancy to allow for more tests to be included in each study period.

VE after a second dose against symptomatic disease with the Omicron variant was also similar to earlier studies (15, 23). Notably, though, VE after the third dose remained high even at 15 or more weeks after vaccination, with limited waning observed. This contrasts with our earlier studies which found substantial waning after a booster dose against symptomatic disease with Omicron. We further investigated this by estimating VE in all women of child-bearing age who did not contribute a test as part of the VE analysis of pregnant women. In these women we found VE waned at 15 or more weeks after the booster, as we have previously reported for the general population (29). While it is unclear why VE against mild disease with Omicron following a third dose appears higher in pregnant than non-pregnant women, it is reassuring that the VE is not lower during pregnancy and such differences may be due to confounding. There were insufficient data to estimate VE against hospitalisation with Omicron in pregnant women as Omicron became dominant towards the end of the study period and all community testing for SARS-CoV-2 stopped in at the end of March 2022.

In addition to protection against mild and severe disease in the mother, maternal vaccination during pregnancy conferred protection in the infant. Both prior infection and maternal vaccination protected infants after birth from symptomatic disease and hospitalisation with Delta and Omicron. In the US, a test-negative, case-control study found 52% (95%CI, 33-65%) VE after two maternal doses against COVID-19 hospitalization overall and 38% (95% C.I.; 8 to 58%) during the Omicron period, increasing to 58% when second dose was administered after 20 weeks gestation (17). In a Canadian study, maternal VE after three doses was 73% (95%CI, 61-80%) against Omicron in infants and 80% (95%CI, 64-89%) against infant hospital admission due to Omicron (18). Similar to the US study, we also found higher protection in the infant when the mother was vaccinated later in pregnancy, with the highest protection after vaccination in the third trimester. There were insufficient data for longer term follow-up infants infected during the Delta period because of the emergence of Omicron but follow-up of infants during the Omicron period indicated that waning of protection does occur in the infant because the level of protection after maternal vaccination in the third trimester was lower at 6-8 months of age than in infants aged 0-5 months. Notably, maternal vaccination after giving birth also had a protective effect in the infant; this could be due to maternal transfer of antibodies in breast-milk (30, 31) as well as lowering her own risk of infection and, therefore, protecting her from infecting her infant.

Prior infection in the mother was also protective against mild and severe disease with Delta and Omicron in the infant. Previous infection with any variant provided high protection in the infant against symptomatic disease and hospitalisation with Delta in the infant, although differential protection by variant was observed - the highest protection was from maternal Delta infection against a subsequent Delta infection in the infant. A protective effect was observed even if the mother’s most recent infection was with wild-type or Alpha against a subsequent Delta infection in her infant, despite these infections occurring many months before the woman became pregnant. The long-lasting duration of protection from previous infection is consistent with other studies (32) but shown here for the first time to extend to infants after birth. Infection with all pre-Omicron variants offered substantially less protection against subsequent mild or severe Omicron infection in the infant, which is known to be more immune evasive than previous variants irrespective of prior infection or vaccination status (32, 33). There were insufficient data to investigate the protection conferred by Omicron infection in the mother against subsequent Omicron disease in the infant. Maternal vaccination achieved greater protection against Omicron disease in the infant than previous infection in the mother, highlighting the importance of maternal vaccination in the Omicron era.

Our study is the largest study of the effectiveness of maternal COVID-19 vaccines on both maternal and infant health. The TNCC design has been well validated and has been widely used for evaluation of COVID-19 vaccines. One of the key strengths is that this approach helps to address unmeasured confounders related to differences in health seeking behaviours and infectious disease exposure between vaccinated and unvaccinated individuals. Nevertheless, there are a number of limitations. This is an observational study that relies on hospital coding – coding errors could result in misclassification in both covariates and outcomes. Similarly, given the observational nature of the study, there may be unmeasured confounders that we were unable to adjust for. Despite the large study, given that the outcomes are rare, there was still significant uncertainty in some of our outcomes, for example, when breaking down the analysis by trimester of vaccination and age of the infant in months.

Together with earlier work from others on the safety of COVID-19 vaccination during pregnancy (1, 2, 8-11), this study adds to a consistent and growing body of evidence of the benefits of maternal vaccination in preventing both mild and severe disease in pregnant women, as well as protection their infants against mild and severe disease during the first six months of life. The findings here support the promotion of both primary and booster vaccination in pregnant women, irrespective of their prior infection status, to protect themselves and their infants.

## Data Availability

All data produced in the present work are contained in the manuscript in aggregate.

## Authors’ Contributions

FCMK and HC wrote the manuscript. JLB, NA, FCMK and MR conceptualised the study. AAM, FCMK and JS curated the data. FCMK and NA conducted the formal analysis. FCMK, NA, AAM and JS accessed and verified the data. All co-authors reviewed the manuscript and were responsible for the decision to submit the manuscript.

## Data Sharing Statement

This work is carried out under Regulation 3 of The Health Service (Control of Patient Information; Secretary of State for Health, 2002) using patient identification information without individual patient consent as part of the UKHSA legal requirement for public health surveillance and monitoring of vaccines. As such, authors cannot make the underlying dataset publicly available for ethical and legal reasons. However, all the data used for this analysis is included as aggregated data in the manuscript tables and appendix. Applications for relevant anonymised data should be submitted to the UKHSA Office for Data Release at https://www.gov.uk/government/publications/accessing-ukhsa-protected-data.

## Ethics Committee Approval

UKHSA has legal permission, provided by Regulation 3 of The Health Service (Control of Patient Information) Regulations 2002, to process patient confidential information for national surveillance of communicable diseases and as such, individual patient consent is not required to access records.

