## Supplementary Appendix for "Vaccine effectiveness against mild and severe disease in pregnant mothers and their infants in England"

### Data Sources

### Hospital Episode Statistics

Data on all women who delivered in 2021 in England were extracted from the Hospital Episode Statistics (HES) (Supplementary Table 1) in November 2022.

### Maternity Services Data Set

Women who delivered in 2021 were linked to the Maternal Services Data Set (MSDS) (Supplementary Table 1) on National Health Service (NHS) number and delivery date to extract the last menstrual period date (LMP).

Data on infants born in 2021, and their mothers, were extracted from the MSDS in November 2022 (Supplementary Table 1). Pregnancy start date was calculated as the earliest possible start date where the gestational age of the infant was known. Where the gestational age was unknown, the date of the last menstrual period (LMP) was used. Where gestational age and LMP were unknown, the earliest possible pregnancy start date was estimated based on the delivery date and whether the infant was term or pre-term. Where it was unknown whether an infant was born term or pre-term, the pregnancy start date was estimated assuming the infant was born at term.

Trimesters were defined as pre-pregnancy, trimester 1 (week 0 (+0 days) to 11 (+6 days)), trimester 2 (week 12 (+0 days) to 26 (+6 days), trimester 3 (week 27 (+0 days) to 15 days before birth (infant only)).

### COVID-19 Testing Data

In the final dataset, only Pillar 1 and 2 tests after 25 November 2020 were included, and Pillar 2 tests were only included up to 31 March 2022, when all community testing was stopped in England. Tests were linked back to all historic positive testing data to identify the date of the most recent positive test (excluding tests within 90 days) prior.

Testing data included all positive PCR and lateral flow tests (LFTs), and negative PCR tests. Any negative tests taken within 7 days of a previous negative test, and any negative tests where symptom onset date was within the 10 days or a previous symptoms onset date for a negative test were dropped as these likely represent the same episode. Negative tests taken within 21 days of a subsequent positive test were also excluded as chances are high that these are false negatives. Positive and negative tests within 90 days of a previous positive test were also excluded; however, where participants had later positive tests within 14 days of a positive then preference was given to PCR tests over LFTs, and tests from symptomatic individuals. To estimate VE against symptomatic disease in pregnant women, data were restricted to Pillar 2 tests from persons who had reported symptoms and gave a symptom onset date within the 10 days before testing to account for reduced PCR sensitivity beyond this period in an infection event. To estimate maternal VE against symptomatic disease in infants, Pillar 1 data were also included. Testing in days 0-5 of life was only in Pillar 1 and almost always negative – reflecting testing in hospital at birth. Only infants aged 6 days to 8 months at symptom onset or date of test were retained.

Past infection in the mother was defined as an infection at least 14 days before birth and was assigned to variant based on the date of the last positive test (in the period until 14 days before birth). Wild type was up until 6 December 2020, Alpha was 7 December 2020 to 9 May 2021, Delta was 10 May 2021 to 12 December 2021 and Omicron was 13 December 2021 onwards (1).

To compare VE in pregnant women with VE in their non-pregnant peers, Pillar 2 testing data were extracted for all women aged 20-44 years. Tests included in the analysis to estimate VE in pregnant women were excluded. A random proxy delivery date between 01 October 2021 and 31 December 2021 was generated for each non-pregnant woman and tests which were 3 months after the delivery date dropped to give this group a similar profile in terms of the period of the test to the pregnant women.

### National Immunisation Management System (NIMS)

The National Immunisation Management System (NIMS) is a national vaccine register containing demographic information on the whole population of England registered with a GP. It is used to record all COVID-19 vaccinations. Testing data were linked to NIMS using combinations of the unique individual NHS number, date of birth, surname, first name, and postcode using deterministic linkage. NIMS was accessed for dates of vaccination and manufacturer, sex, date of birth, ethnicity, and residential address. Addresses were used to determine index of multiple deprivation quintile. Data on risk group status, clinically extremely vulnerable status, severely immunosuppressed and health/social care worker status were also extracted from the NIMS.

Booster doses were identified as a third dose given at least 84 days after a second dose and administered after 13 September 2021. Individuals with four or more doses of vaccine, heterologous primary schedule or fewer than 19 days between their first and second dose were excluded.

Only tests from women who could be linked to the NIMS and tests from infants whose mother could be linked to the NIMS were included. Tests from pregnant women who received anything other than a ChAdOx1-S, BNT162b2 or mRNA1273 primary course and other than a BNT162b2 or mRNA1273 booster dose were excluded. Infants were also excluded if the mother was given a vaccine other than ChAdOx1-S, BNT162b2 or mRNA1273, if the infant had received a vaccine prior to onset date and if the week of birth was prior to 2 May 2021 as there was minimal maternal vaccination prior to this date, and only women at high risk of exposure or severe disease were vaccinated before mid-April 2021.

Hospital Admission Data

Hospital inpatient admissions for a range of acute respiratory illnesses were identified from the Secondary Uses Service (SUS) (2) (Supplementary Table 1) and were linked to the testing data in November 2022 using NHS number and date of birth. Admissions with an ICD-10 coded acute respiratory illness (ARI) discharge diagnosis (Supplementary Table 2) in any diagnosis field were identified where the sample was taken 1 day before and up to 2 days after the admission. Data were restricted to those with ARI in the first diagnosis field and where the length of stay was at least two days.

### Identification of Delta and Omicron Variants and assignment to cases

Cases were defined as Delta or Omicron (BA.1 and BA.2) based on whole genome sequencing, genotyping, S-gene target failure (SGTF) status or period, with sequencing taking priority, followed by genotyping, SGTF status and period (Supplementary Table 1). Where subsequent positive tests within 14 days included sequencing, genotyping or S-gene target failure information, this information was used to classify the variant. The Delta analysis was restricted from 26 April 2021 to 9 January 2022. The Omicron analysis was restricted from 29 November 2021 onwards. Previously we have not found a difference in VE between BA.1 and BA.2 (24) so these were both included in the Omicron analyses.

Supplementary Table 1. Description of data sources used to estimate vaccine effectiveness. Women who were delivered in 2021 were identified from HES and infants born in 2021 were identified from MSDS. Cases (those testing positive) and controls (those testing negative) were identified in the testing data. Data were linked to the NIMS to ascertain vaccination status, to the genomics line list to ascertain variant information, and to the SUS hospitalisation records.

| Data Source | Description | Variables |
| --- | --- | --- |
| HES | Data on all women who delivered in 2021 in England were extracted from the HES. | Delivery date, gestational age of the infant, information on whether the infant was term or pre-term, the earliest and latest possible pregnancy start dates, last menstrual period date (LMPD). |
| MSDS | Data on infants born in 2021, and their mothers, were extracted from the MSDS. | NHS number of the infant and mother, LPMD, delivery date, gestational age of the infant, information on whether the infant was term or pre-term and the pregnancy start date. |
| Pillar 1 and 2 testing data | SARS-CoV-2 PCR testing in England is undertaken by hospital and public health laboratories (Pillar 1), as well as by community testing (Pillar 2). Pillar 1 testing data is available from UKHSA labs and NHS hospitals for those with a clinical need, and health and care workers. Pillar 2 community testing is available to anyone with symptoms consistent with COVID-19 (high temperature, new continuous cough, or loss or change in sense of smell or taste), anyone who is a contact of a confirmed case, care home staff and residents, and to those who have self-tested as positive using a lateral flow test (LFT). SGTF information to classify variant is available from some Pillar 2 tests. | Date of birth (age), period (week of test), S gene target failure (SGTF) status, previous positivity |
| NIMS vaccination record | The National Immunization Management System (NIMS) contains demographic information on the whole population of England who are registered with a general practice physician in England and is used to record all COVID-19 vaccinations. | Infant and maternal vaccination dates by manufacturer, index of multiple deprivation, ethnicity, geographic region, date of birth, health and social care worker status, clinical risk group status, clinically extremely vulnerable, severely immunosuppressed. Clinical risk groups included a range of chronic conditions as described in the Green Book (3), whereas the CEV group included persons who were considered to be at the highest risk for severe COVID-19, including those with immunosuppressed conditions and those with severe respiratory disease. The CEV flag is a record of vulnerable patients thought to be at high risk of complications from COVID-19. The severely immunosuppressed flag is also provided by NHS Digital and includes those with specific immunosuppression conditions. The severely immunosuppressed were offered additional doses as part of the primary vaccination schedule which the CEV were not. |
| Genomics data | Sequencing and genotyping is undertaken through a network of laboratories, including the Wellcome Sanger Institute. Whole-genome sequences are assigned to UKHSA definitions of variants based on mutations. Data were provided by the Genomics Cell at UKHSA. | Whole genome sequencing and genotyping data associated with Pillar 1 and 2 testing data. |
| Secondary Care Hospital Admission Data (SUS) | SUS is the national electronic database of hospital admissions that provides timely updates of ICD-10 codes for completed hospital stays for all NHS hospitals in England. Up to 24 ICD-10 diagnoses fields can be completed in SUS for each admission with the first diagnosis field indicating the primary reason for admission. Length of stay was calculated as date of discharge – date of admission. Where multiple admissions linked to the same sample date the first admission after the sample date was retained and episode length calculated by summing the stay length for each admission. Data were restricted to those with ARI in the first diagnosis field and where the length of stay was at least two days. | Primary diagnosis code, length of stay, treatment with oxygen, mechanical ventilation or treatment on intensive care unit (ICU). |

Supplementary Table 2. SUS Acute respiratory illness ICD10 code list.

| SUS Acute respiratory illness ICD10 code list | |
| --- | --- |
| J04* | Acute laryngitis and tracheitis |
| J09* | Influenza due to identified avian influenza virus |
| J10* | Influenza with pneumonia, other influenza virus identified |
| J11* | Influenza with pneumonia, virus not identified |
| J12* | Viral pneumonia, not elsewhere classified |
| J13* | Pneumonia due to Streptococcus pneumoniae |
| J14* | Pneumonia due to Haemophilus influenzae |
| J15* | Bacterial pneumonia, not elsewhere classified |
| J16* | Pneumonia due to other infectious organisms, not elsewhere classified |
| J17* | Pneumonia in diseases classified elsewhere |
| J18* | Pneumonia, organism unspecified |
| J20* | Acute bronchitis |
| J21* | Acute bronchiolitis |
| J22* | Unspecified acute lower respiratory infection |
| J80* | ARDS (related to respiratory infection) |
| U071, U072 | COVID-19, virus identified and not identified |
| U04* | Severe acute respiratory syndrome (SARS) |

Supplementary Table 3. Descriptive characteristics of tests from symptomatic women who delivered in 2021 and for whom VE was estimated in Table 1 and Figure 1.

|  |  |  | **Delta period** | | | | **Omicron period** | | | |
| --- | --- | --- | --- | --- | --- | --- | --- | --- | --- | --- |
|  |  |  | **Control** | | **Case** | | **Control** | | **Case** | |
|  |  |  | **n** | **%** | **n** | **%** | **n** | **%** | **n** | **%** |
|  | **Test Result** | **Interval (weeks)** | **35,210** | **67.8%** | **16,693** | **32.2%** | **5,974** | **55.9%** | **4,715** | **44.1%** |
| **Vaccination Status** | Unvaccinated |  | 17,013 | **48.3%** | 11,442 | **68.5%** | 1,290 | **21.6%** | 1,773 | **37.6%** |
|  | Dose 1 | 0-1 | 1,467 | **4.2%** | 648 | **3.9%** | 160 | **2.7%** | 138 | **2.9%** |
|  |  | 2-4 | 2,538 | **7.2%** | 761 | **4.6%** | 256 | **4.3%** | 259 | **5.5%** |
|  |  | 5-9 | 2,742 | **7.8%** | 1,162 | **7.0%** | 303 | **5.1%** | 271 | **5.7%** |
|  |  | 10-14 | 319 | **0.9%** | 103 | **0.6%** | 34 | **0.6%** | 27 | **0.6%** |
|  |  | 15-19 | 325 | **0.9%** | 92 | **0.6%** | 33 | **0.6%** | 17 | **0.4%** |
|  |  | 20-24 | 277 | **0.8%** | 168 | **1.0%** | 17 | **0.3%** | 12 | **0.3%** |
|  |  | 25+ | 770 | **2.2%** | 494 | **3.0%** | 190 | **3.2%** | 202 | **4.3%** |
|  | Dose 2 | 0-1 | 1,098 | **3.1%** | 272 | **1.6%** | 152 | **2.5%** | 92 | **2.0%** |
|  |  | 2-4 | 1,332 | **3.8%** | 149 | **0.9%** | 165 | **2.8%** | 99 | **2.1%** |
|  |  | 5-9 | 1,990 | **5.7%** | 305 | **1.8%** | 340 | **5.7%** | 150 | **3.2%** |
|  |  | 10-14 | 1,724 | **4.9%** | 363 | **2.2%** | 421 | **7.0%** | 244 | **5.2%** |
|  |  | 15-19 | 1,335 | **3.8%** | 334 | **2.0%** | 567 | **9.5%** | 217 | **4.6%** |
|  |  | 20-24 | 737 | **2.1%** | 211 | **1.3%** | 323 | **5.4%** | 151 | **3.2%** |
|  |  | 25+ | 544 | **1.5%** | 160 | **1.0%** | 245 | **4.1%** | 214 | **4.5%** |
|  | Dose 3 | 0-1 | 575 | **1.6%** | 20 | **0.1%** | 557 | **9.3%** | 297 | **6.3%** |
|  |  | 2-4 | 243 | **0.7%** | 7 | **0.0%** | 410 | **6.9%** | 236 | **5.0%** |
|  |  | 5-9 | 131 | **0.4%** | 2 | **0.0%** | 346 | **5.8%** | 189 | **4.0%** |
|  |  | 10-14 | 50 | **0.1%** | 0 | **0.0%** | 113 | **1.9%** | 89 | **1.9%** |
|  |  | 15+ | 0 | **0.0%** | 0 | **0.0%** | 52 | **0.9%** | 38 | **0.8%** |
| **Age** | 16-19 |  | 541 | **1.5%** | 408 | **2.4%** | 109 | **1.8%** | 107 | **2.3%** |
|  | 20-24 |  | 3,347 | **9.5%** | 2,325 | **13.9%** | 530 | **8.9%** | 632 | **13.4%** |
|  | 25-29 |  | 8,746 | **24.8%** | 4,822 | **28.9%** | 1,403 | **23.5%** | 1,423 | **30.2%** |
|  | 30-34 |  | 13,496 | **38.3%** | 5,617 | **33.6%** | 2,349 | **39.3%** | 1,592 | **33.8%** |
|  | 35-39 |  | 7,620 | **21.6%** | 2,851 | **17.1%** | 1,325 | **22.2%** | 775 | **16.4%** |
|  | 40-44 |  | 1,413 | **4.0%** | 648 | **3.9%** | 249 | **4.2%** | 177 | **3.8%** |
|  | 45-49 |  | 47 | **0.1%** | 21 | **0.1%** | 9 | **0.2%** | 9 | **0.2%** |
|  | 50-54 |  | 0 | **0.0%** | 1 | **0.0%** | 0 | **0.0%** | 0 | **0.0%** |
| **Ethnicity** | African |  | 367 | **1.0%** | 199 | **1.2%** | 50 | **0.8%** | 68 | **1.4%** |
|  | Any other Asian background | | 655 | **1.9%** | 240 | **1.4%** | 114 | **1.9%** | 90 | **1.9%** |
|  | Any other Black background | | 139 | **0.4%** | 119 | **0.7%** | 17 | **0.3%** | 27 | **0.6%** |
|  | Any other White background | | 2,952 | **8.4%** | 1,549 | **9.3%** | 496 | **8.3%** | 590 | **12.5%** |
|  | Any other ethnic group |  | 731 | **2.1%** | 354 | **2.1%** | 99 | **1.7%** | 113 | **2.4%** |
|  | Any other mixed background | | 240 | **0.7%** | 122 | **0.7%** | 52 | **0.9%** | 44 | **0.9%** |
|  | Bangladeshi or British Bangladeshi | | 371 | **1.1%** | 195 | **1.2%** | 41 | **0.7%** | 52 | **1.1%** |
|  | British, Mixed British |  | 25,016 | **71.0%** | 11,887 | **71.2%** | 4,393 | **73.5%** | 3,143 | **66.7%** |
|  | Caribbean |  | 114 | **0.3%** | 103 | **0.6%** | 17 | **0.3%** | 29 | **0.6%** |
|  | Chinese |  | 203 | **0.6%** | 46 | **0.3%** | 38 | **0.6%** | 21 | **0.4%** |
|  | Indian or British Indian |  | 1,280 | **3.6%** | 379 | **2.3%** | 196 | **3.3%** | 115 | **2.4%** |
|  | Irish |  | 254 | **0.7%** | 71 | **0.4%** | 45 | **0.8%** | 28 | **0.6%** |
|  | Pakistani or British Pakistani | | 1,090 | **3.1%** | 603 | **3.6%** | 133 | **2.2%** | 128 | **2.7%** |
|  | White and Asian |  | 128 | **0.4%** | 49 | **0.3%** | 19 | **0.3%** | 22 | **0.5%** |
|  | White and Black African |  | 84 | **0.2%** | 35 | **0.2%** | 15 | **0.3%** | 20 | **0.4%** |
|  | White and Black Caribbean | | 188 | **0.5%** | 122 | **0.7%** | 28 | **0.5%** | 26 | **0.6%** |
|  | Missing |  | 1,398 | **4.0%** | 620 | **3.7%** | 221 | **3.7%** | 199 | **4.2%** |
| **NHS Region** | East of England |  | 4,217 | **12.0%** | 1,672 | **10.0%** | 848 | **14.2%** | 515 | **10.9%** |
|  | London |  | 4,756 | **13.5%** | 1,763 | **10.6%** | 864 | **14.5%** | 687 | **14.6%** |
|  | Midlands |  | 6,270 | **17.8%** | 3,477 | **20.8%** | 970 | **16.2%** | 885 | **18.8%** |
|  | North East |  | 5,517 | **15.7%** | 3,359 | **20.1%** | 821 | **13.7%** | 870 | **18.5%** |
|  | North West |  | 4,632 | **13.2%** | 2,834 | **17.0%** | 692 | **11.6%** | 732 | **15.5%** |
|  | South East |  | 5,782 | **16.4%** | 2,096 | **12.6%** | 1,078 | **18.0%** | 683 | **14.5%** |
|  | South West |  | 4,036 | **11.5%** | 1,492 | **8.9%** | 701 | **11.7%** | 343 | **7.3%** |
|  | Missing |  | 0 | **0.0%** | 0 | **0.0%** | 0 | **0.0%** | 0 | **0.0%** |
| **IMD Quintiles** | 1 |  | 6,481 | **18.4%** | 4,438 | **26.6%** | 895 | **15.0%** | 1,130 | **24.0%** |
|  | 2 |  | 6,527 | **18.5%** | 3,583 | **21.5%** | 1,099 | **18.4%** | 972 | **20.6%** |
|  | 3 |  | 7,049 | **20.0%** | 3,226 | **19.3%** | 1,266 | **21.2%** | 925 | **19.6%** |
|  | 4 |  | 7,284 | **20.7%** | 2,894 | **17.3%** | 1,282 | **21.5%** | 881 | **18.7%** |
|  | 5 |  | 7,631 | **21.7%** | 2,432 | **14.6%** | 1,389 | **23.3%** | 770 | **16.3%** |
|  | Missing |  | 238 | **0.7%** | 120 | **0.7%** | 43 | **0.7%** | 37 | **0.8%** |
| **Vaccine priority groups** | HSCW |  | 3,686 | **10.5%** | 975 | **5.8%** | 664 | **11.1%** | 379 | **8.0%** |
|  | At risk |  | 6,349 | **18.0%** | 2,932 | **17.6%** | 1,086 | **18.2%** | 838 | **17.8%** |
|  | Severely Immunosuppressed | | 90 | **0.3%** | 45 | **0.3%** | 19 | **0.3%** | 8 | **0.2%** |
|  | CEV |  | 1,776 | **5.0%** | 713 | **4.3%** | 264 | **4.4%** | 186 | **3.9%** |
| **Variant of most recent previous infection** | None |  | 31,595 | **89.7%** | 16,371 | **98.1%** | 5,038 | **84.3%** | 4,204 | **89.2%** |
|  | Wild-type |  | 1,332 | **3.8%** | 170 | **1.0%** | 242 | **4.1%** | 146 | **3.1%** |
|  | Alpha |  | 1,876 | **5.3%** | 140 | **0.8%** | 356 | **6.0%** | 189 | **4.0%** |
|  | Delta |  | 407 | **1.2%** | 12 | **0.1%** | 334 | **5.6%** | 176 | **3.7%** |
|  | Omicron |  | 0 | **0.0%** | 0 | **0.0%** | 4 | **0.1%** | 0 | **0.0%** |

Supplementary Table 4. Vaccine effectiveness against symptomatic disease with Delta for pregnant women, only including tests from pregnancy start date up to the delivery date.

|  |  | Delta | | | |
| --- | --- | --- | --- | --- | --- |
| Vaccination Status | Interval (weeks) | Cases | Controls | OR (95% CI) | VE (95% CI) |
| Unvaccinated |  | 7,492 | 12,002 | Baseline | Baseline |
| Dose 1* | 0-1 | 210 | 624 | 0.54 (0.46-0.65) | 45.7 (35.5 to 54.3) |
|  | 2-4 | 288 | 1,056 | 0.39 (0.34-0.45) | 60.5 (54.6 to 65.7) |
|  | 5-9 | 470 | 1,143 | 0.49 (0.43-0.55) | 51.1 (45.0 to 56.6) |
|  | 10-14 | 50 | 222 | 0.41 (0.29-0.57) | 59.3 (42.9 to 71.0) |
|  | 15-19 | 80 | 274 | 0.46 (0.35-0.6) | 54.2 (39.8 to 65.2) |
|  | 20-24 | 157 | 256 | 0.72 (0.58-0.89) | 28.4 (11.2 to 42.3) |
|  | 25+ | 416 | 597 | 0.81 (0.7-0.93) | 19.4 (7.1 to 30.0) |
| Dose 2* | 0-1 | 155 | 556 | 0.31 (0.25-0.37) | 69.2 (62.8 to 74.5) |
|  | 2-4 | 108 | 796 | 0.15 (0.12-0.18) | 85.1 (81.6 to 88.0) |
|  | 5-9 | 207 | 1,190 | 0.22 (0.19-0.26) | 78.0 (74.2 to 81.2) |
|  | 10-14 | 223 | 851 | 0.3 (0.26-0.35) | 70.0 (64.8 to 74.5) |
|  | 15-19 | 193 | 532 | 0.41 (0.34-0.49) | 58.7 (50.6 to 65.5) |
|  | 20-24 | 138 | 366 | 0.45 (0.36-0.55) | 55.3 (44.7 to 63.8) |
|  | 25+ | 103 | 320 | 0.43 (0.33-0.54) | 57.3 (45.5 to 66.6) |
| Dose 3** | 0-1 | 5 | 77 | 0.1 (0.04-0.24) | 90.5 (76.0 to 96.2) |
|  | 2-4 | 3 | 62 | 0.06 (0.02-0.19) | 94.0 (80.8 to 98.1) |
|  | 5-9 | 1 | 47 | 0.03 (0-0.21) | 97.1 (78.9 to 99.6) |
|  | 10-14 | 0 | 1 | n too small | n too small |
|  | 15+ | 0 | 0 | n too small | n too small |
| *Primary course manufacturer ChAdOx1-S, BNT162b2 or mRNA1273 | | | | |  |
| **Booster manufacturer BNT162b2 or mRNA1273 | | | |  |  |

Supplementary Table 5. Vaccine effectiveness against symptomatic disease with Omicron for non-pregnant women aged 20 to 44 in the same study period.

|  |  | Omicron | | | |
| --- | --- | --- | --- | --- | --- |
| Vaccination Status | Interval (weeks) | Cases | Controls | OR (95% CI) | VE (95% CI) |
| Unvaccinated |  | 86,441 | 55,783 | Baseline | Baseline |
| Dose 1* | 0-1 | 2,390 | 2,199 | 0.66 (0.62-0.71) | 33.8 (29.3 to 38.0) |
|  | 2-4 | 2,975 | 3,385 | 0.49 (0.46-0.52) | 50.9 (48.1 to 53.6) |
|  | 5-9 | 4,191 | 4,458 | 0.57 (0.55-0.6) | 42.7 (39.9 to 45.5) |
|  | 10-14 | 1,757 | 2,050 | 0.6 (0.56-0.65) | 40.1 (35.5 to 44.4) |
|  | 15-19 | 1,776 | 1,947 | 0.74 (0.69-0.8) | 25.5 (19.6 to 31) |
|  | 20-24 | 2,542 | 3,003 | 0.76 (0.71-0.81) | 24.0 (19 to 28.7) |
|  | 25+ | 9,550 | 8,152 | 0.73 (0.71-0.76) | 26.9 (24.3 to 29.5) |
| Dose 2* | 0-1 | 2,085 | 3,326 | 0.43 (0.4-0.45) | 57.4 (54.7 to 60.0) |
|  | 2-4 | 3,463 | 5,984 | 0.37 (0.35-0.39) | 63.2 (61.3 to 64.9) |
|  | 5-9 | 9,081 | 12,789 | 0.53 (0.51-0.54) | 47.5 (45.7 to 49.2) |
|  | 10-14 | 19,551 | 34,442 | 0.65 (0.64-0.67) | 34.7 (33.1 to 36.3) |
|  | 15-19 | 58,747 | 85,901 | 0.77 (0.76-0.79) | 22.9 (21.5 to 24.3) |
|  | 20-24 | 55,656 | 63,750 | 0.82 (0.8-0.83) | 18.1 (16.6 to 19.6) |
|  | 25+ | 92,014 | 82,758 | 0.93 (0.92-0.95) | 6.5 (4.9 to 8.1) |
| Dose 3** | 0-1 | 55,324 | 73,073 | 0.47 (0.46-0.48) | 53.3 (52.4 to 54.1) |
|  | 2-4 | 59,261 | 80,388 | 0.33 (0.33-0.34) | 66.9 (66.3 to 67.4) |
|  | 5-9 | 61,381 | 83,411 | 0.41 (0.4-0.42) | 59.2 (58.5 to 59.9) |
|  | 10-14 | 37,961 | 32,880 | 0.53 (0.52-0.54) | 46.8 (45.6 to 47.9) |
|  | 15+ | 12,787 | 8,396 | 0.68 (0.65-0.7) | 32.4 (30.1 to 34.7) |
| *Primary course manufacturer ChAdOx1-S, BNT162b2 or mRNA1273 | | | | |  |
| **Booster manufacturer BNT162b2 or mRNA1273 | | | |  |  |

Supplementary Table 6. Descriptive characteristics of tests from hospitalised women who delivered in 2021 and for whom VE was estimated.

|  |  |  | **Delta** | | | |
| --- | --- | --- | --- | --- | --- | --- |
|  |  |  | **Control** | | **Case** | |
|  |  |  | **n** | **%** | **n** | **%** |
|  | **Test Result** | **Interval (weeks)** | **86** | **6.4%** | **1,249** | **93.6%** |
| **Vaccination Status** | Unvaccinated | | 55 | **64.0%** | 1,109 | **88.8%** |
|  | Dose 1 | 0-4 | 4 | **4.7%** | 32 | **2.6%** |
|  |  | 5-19 | 8 | **9.3%** | 26 | **2.1%** |
|  |  | 20+ | 2 | **2.3%** | 30 | **2.4%** |
|  | Dose 2 | 0-4 | 4 | **4.7%** | 4 | **0.3%** |
|  |  | 5-19 | 12 | **14.0%** | 34 | **2.7%** |
|  |  | 20+ | 1 | **1.2%** | 14 | **1.1%** |
| **Age** | 16-19 |  | 2 | **2.3%** | 23 | **1.8%** |
|  | 20-24 |  | 12 | **14.0%** | 139 | **11.1%** |
|  | 25-29 |  | 16 | **18.6%** | 351 | **28.1%** |
|  | 30-34 |  | 41 | **47.7%** | 386 | **30.9%** |
|  | 35-39 |  | 13 | **15.1%** | 275 | **22.0%** |
|  | 40-44 |  | 2 | **2.3%** | 67 | **5.4%** |
|  | 45-49 |  | 0 | **0.0%** | 4 | **0.3%** |
|  | 50-54 |  | 0 | **0.0%** | 4 | **0.3%** |
| **Ethnicity** | African |  | 2 | **2.3%** | 51 | **4.1%** |
|  | Any other Asian background | | 1 | **1.2%** | 35 | **2.8%** |
|  | Any other Black background | | 0 | **0.0%** | 22 | **1.8%** |
|  | Any other White background | | 7 | **8.1%** | 154 | **12.3%** |
|  | Any other ethnic group | | 2 | **2.3%** | 63 | **5.0%** |
|  | Any other mixed background | | 0 | **0.0%** | 9 | **0.7%** |
|  | Bangladeshi or British Bangladeshi | | 2 | **2.3%** | 32 | **2.6%** |
|  | British, Mixed British | | 56 | **65.1%** | 650 | **52.0%** |
|  | Caribbean |  | 0 | **0.0%** | 26 | **2.1%** |
|  | Chinese |  | 0 | **0.0%** | 11 | **0.9%** |
|  | Indian or British Indian | | 6 | **7.0%** | 27 | **2.2%** |
|  | Irish |  | 0 | **0.0%** | 6 | **0.5%** |
|  | Pakistani or British Pakistani | | 7 | **8.1%** | 95 | **7.6%** |
|  | White and Asian | | 0 | **0.0%** | 11 | **0.9%** |
|  | White and Black African | | 0 | **0.0%** | 4 | **0.3%** |
|  | White and Black Caribbean | | 1 | **1.2%** | 12 | **1.0%** |
|  | Missing |  | 2 | **2.3%** | 41 | **3.3%** |
| **NHS Region** | East of England | | 11 | **12.8%** | 138 | **11.0%** |
|  | London |  | 9 | **10.5%** | 233 | **18.7%** |
|  | Midlands |  | 18 | **20.9%** | 355 | **28.4%** |
|  | North East |  | 18 | **20.9%** | 177 | **14.2%** |
|  | North West |  | 13 | **15.1%** | 176 | **14.1%** |
|  | South East |  | 12 | **14.0%** | 107 | **8.6%** |
|  | South West |  | 5 | **5.8%** | 63 | **5.0%** |
|  | Missing |  | 0 | **0.0%** | 0 | **0.0%** |
| **IMD Quintiles** | 1 |  | 17 | **19.8%** | 422 | **33.8%** |
|  | 2 |  | 23 | **26.7%** | 284 | **22.7%** |
|  | 3 |  | 17 | **19.8%** | 221 | **17.7%** |
|  | 4 |  | 19 | **22.1%** | 168 | **13.5%** |
|  | 5 |  | 10 | **11.6%** | 149 | **11.9%** |
|  | Missing |  | 0 | **0.0%** | 5 | **0.4%** |
| **Vaccine priority groups** | HSCW |  | 10 | **11.6%** | 56 | **4.5%** |
|  | At risk |  | 32 | **37.2%** | 299 | **23.9%** |
|  | Severely Immunosuppressed | | 0 | **0.0%** | 2 | **0.2%** |
|  | CEV |  | 6 | **7.0%** | 65 | **5.2%** |
| **Variant of most recent previous infection** | None |  | 78 | **90.7%** | 1,238 | **99.1%** |
|  | Wild-type |  | 1 | **1.2%** | 4 | **0.3%** |
|  | Alpha |  | 5 | **5.8%** | 5 | **0.4%** |
|  | Delta |  | 2 | **2.3%** | 2 | **0.2%** |
|  | Omicron |  | 0 | **0.0%** | 0 | **0.0%** |

Supplementary Table 7. Vaccine effectiveness against hospitalisation with Delta in pregnant women within 3 months of their pregnancy start up to 3 months post-delivery in England.

| Vaccination Status | Interval (weeks) | Cases | Controls | OR (95% CI) | VE (95% CI) |
| --- | --- | --- | --- | --- | --- |
| Unvaccinated |  | 1109 | 55 | Baseline | Baseline |
| Dose 1* | 0-4 | 32 | 4 | 0.32 (0.09-1.07) | 69.2 (-4.1 to 90.9) |
|  | 5-19 | 26 | 8 | 0.06 (0.02-0.19) | 94.1 (81.2 to 98.1) |
|  | 20+ | 30 | 2 | n too small | n too small |
| Dose 2* | 0-4 | 4 | 4 | n too small | n too small |
|  | 5-19 | 34 | 12 | 0.08 (0.03-0.22) | 92.7 (79.9 to 97.4) |
|  | 20+ | 14 | 1 | n too small | n too small |
| *Primary course manufacturer ChAdOx1-S, BNT162b2 or mRNA1273 | | | | |  |

Supplementary Table 8. Descriptive characteristics of tests from symptomatic infants for whom VE was estimated in Figures 2-3.

|  |  |  |  | **Delta period** | | | | **Omicron period** | | | |
| --- | --- | --- | --- | --- | --- | --- | --- | --- | --- | --- | --- |
|  | **Test Result** |  |  | **Control (Negative)** | | **Case (Positive)** | | **Control (Negative)** | | **Case (Positive)** | |
|  |  |  |  | **n** | **%** | **n** | **%** | **n** | **%** | **n** | **%** |
|  |  |  |  | **23,053** | **88.7%** | **2,924** | **11.3%** | **13,908** | **71.0%** | **5,669** | **29.0%** |
| **Pillar** | 1 |  |  | 15,525 | **67.3%** | 1,060 | **36.3%** | 7,007 | **50.4%** | 1,672 | **29.5%** |
|  | 2 |  |  | 7,528 | **32.7%** | 1,864 | **63.7%** | 6,901 | **49.6%** | 3,997 | **70.5%** |
| **Gender of infant** | Female |  |  | 10,108 | **43.8%** | 1,344 | **46.0%** | 6,062 | **43.6%** | 2,614 | **46.1%** |
|  | Male |  |  | 12,847 | **55.7%** | 1,578 | **54.0%** | 7,815 | **56.2%** | 3,052 | **53.8%** |
|  | Unknown |  |  | 98 | **0.4%** | 2 | **0.1%** | 31 | **0.2%** | 3 | **0.1%** |
| **Premature** | No |  |  | 21,121 | **91.6%** | 2,740 | **93.7%** | 12,810 | **92.1%** | 5,280 | **93.1%** |
|  | Gestational age <37 weeks | |  | 1,518 | **6.6%** | 163 | **5.6%** | 782 | **5.6%** | 306 | **5.4%** |
|  | Gestational age <33 weeks | |  | 414 | **1.8%** | 21 | **0.7%** | 316 | **2.3%** | 83 | **1.5%** |
| **Age of infant (months)** | 0 |  |  | 4,481 | **19.4%** | 402 | **13.7%** | 914 | **6.6%** | 156 | **2.8%** |
|  | 1 |  |  | 5,881 | **25.5%** | 784 | **26.8%** | 1,823 | **13.1%** | 577 | **10.2%** |
|  | 2 |  |  | 3,749 | **16.3%** | 577 | **19.7%** | 1,648 | **11.8%** | 691 | **12.2%** |
|  | 3 |  |  | 2,857 | **12.4%** | 414 | **14.2%** | 1,666 | **12.0%** | 684 | **12.1%** |
|  | 4 |  |  | 2,314 | **10.0%** | 347 | **11.9%** | 1,670 | **12.0%** | 757 | **13.4%** |
|  | 5 |  |  | 1,840 | **8.0%** | 237 | **8.1%** | 1,840 | **13.2%** | 856 | **15.1%** |
|  | 6 |  |  | 1,357 | **5.9%** | 137 | **4.7%** | 1,949 | **14.0%** | 810 | **14.3%** |
|  | 7 |  |  | 564 | **2.4%** | 26 | **0.9%** | 1,468 | **10.6%** | 731 | **12.9%** |
|  | 8 |  |  | 10 | **0.0%** | 0 | **0.0%** | 930 | **6.7%** | 407 | **7.2%** |
| **Ethnicity** | African |  |  | 406 | **1.8%** | 42 | **1.4%** | 202 | **1.5%** | 93 | **1.6%** |
|  | Any other Asian background | |  | 412 | **1.8%** | 51 | **1.7%** | 274 | **2.0%** | 113 | **2.0%** |
|  | Any other Black background | |  | 151 | **0.7%** | 18 | **0.6%** | 86 | **0.6%** | 32 | **0.6%** |
|  | Any other White background | |  | 1,441 | **6.3%** | 219 | **7.5%** | 909 | **6.5%** | 608 | **10.7%** |
|  | Any other ethnic group |  |  | 490 | **2.1%** | 71 | **2.4%** | 296 | **2.1%** | 129 | **2.3%** |
|  | Any other mixed background | |  | 461 | **2.0%** | 47 | **1.6%** | 268 | **1.9%** | 125 | **2.2%** |
|  | Bangladeshi or British Bangladeshi | |  | 231 | **1.0%** | 37 | **1.3%** | 116 | **0.8%** | 40 | **0.7%** |
|  | British, Mixed British |  |  | 12,643 | **54.8%** | 1,500 | **51.3%** | 8,218 | **59.1%** | 2,966 | **52.3%** |
|  | Caribbean |  |  | 62 | **0.3%** | 13 | **0.4%** | 35 | **0.3%** | 24 | **0.4%** |
|  | Chinese |  |  | 64 | **0.3%** | 1 | **0.0%** | 32 | **0.2%** | 19 | **0.3%** |
|  | Indian or British Indian |  |  | 537 | **2.3%** | 51 | **1.7%** | 323 | **2.3%** | 180 | **3.2%** |
|  | Irish |  |  | 75 | **0.3%** | 11 | **0.4%** | 51 | **0.4%** | 15 | **0.3%** |
|  | Pakistani or British Pakistani | |  | 664 | **2.9%** | 71 | **2.4%** | 399 | **2.9%** | 186 | **3.3%** |
|  | White and Asian |  |  | 213 | **0.9%** | 26 | **0.9%** | 161 | **1.2%** | 63 | **1.1%** |
|  | White and Black African |  |  | 144 | **0.6%** | 11 | **0.4%** | 98 | **0.7%** | 46 | **0.8%** |
|  | White and Black Caribbean | |  | 215 | **0.9%** | 37 | **1.3%** | 133 | **1.0%** | 63 | **1.1%** |
|  | Missing |  |  | 4,844 | **21.0%** | 718 | **24.6%** | 2,307 | **16.6%** | 967 | **17.1%** |
| **IMD Quintiles** | 1 |  |  | 5,562 | **24.1%** | 818 | **28.0%** | 3,112 | **22.4%** | 1,388 | **24.5%** |
|  | 2 |  |  | 4,437 | **19.2%** | 585 | **20.0%** | 2,670 | **19.2%** | 1,127 | **19.9%** |
|  | 3 |  |  | 4,319 | **18.7%** | 575 | **19.7%** | 2,647 | **19.0%** | 1,087 | **19.2%** |
|  | 4 |  |  | 4,258 | **18.5%** | 497 | **17.0%** | 2,672 | **19.2%** | 1,068 | **18.8%** |
|  | 5 |  |  | 4,247 | **18.4%** | 413 | **14.1%** | 2,671 | **19.2%** | 945 | **16.7%** |
|  | Missing |  |  | 230 | **1.0%** | 36 | **1.2%** | 136 | **1.0%** | 54 | **1.0%** |
| **Age of mother (years)** | 15-24 |  |  | 3,510 | **15.2%** | 509 | **17.4%** | 1,933 | **13.9%** | 898 | **15.8%** |
|  | 25-29 |  |  | 6,308 | **27.4%** | 758 | **25.9%** | 3,579 | **25.7%** | 1,488 | **26.2%** |
|  | 30-34 |  |  | 7,965 | **34.6%** | 963 | **32.9%** | 4,953 | **35.6%** | 1,934 | **34.1%** |
|  | 35+ |  |  | 5,270 | **22.9%** | 694 | **23.7%** | 3,443 | **24.8%** | 1,349 | **23.8%** |
| **Mother's vaccination status; no. of doses in pregnancy and no. of doses post pregnancy** | Unvaccinated | 0 | 0 | 10,452 | **45.3%** | 1,648 | **56.4%** | 3,898 | **28.0%** | 1,780 | **31.4%** |
|  | Unvaccinated | 0 | 1 | 3,772 | **16.4%** | 572 | **19.6%** | 1,367 | **9.8%** | 652 | **11.5%** |
|  | Unvaccinated | 0 | 2 | 3,842 | **16.7%** | 444 | **15.2%** | 4,028 | **29.0%** | 1,736 | **30.6%** |
|  | Pre-pregnancy | 1 | 0 | 62 | **0.3%** | 3 | **0.1%** | 95 | **0.7%** | 39 | **0.7%** |
|  | Pre-pregnancy | 1 | 1 | 12 | **0.1%** | 0 | **0.0%** | 48 | **0.3%** | 21 | **0.4%** |
|  | Pre-pregnancy | 2 | 0 | 10 | **0.0%** | 0 | **0.0%** | 21 | **0.2%** | 6 | **0.1%** |
|  | Trimester 1 | 1 | 0 | 144 | **0.6%** | 15 | **0.5%** | 122 | **0.9%** | 48 | **0.8%** |
|  | Trimester 1 | 1 | 1 | 38 | **0.2%** | 2 | **0.1%** | 85 | **0.6%** | 39 | **0.7%** |
|  | Trimester 1 | 2 | 0 | 70 | **0.3%** | 3 | **0.1%** | 105 | **0.8%** | 40 | **0.7%** |
|  | Trimester 1 | 2 | 1 | 19 | **0.1%** | 2 | **0.1%** | 99 | **0.7%** | 37 | **0.7%** |
|  | Trimester 2 | 1 | 0 | 69 | **0.3%** | 9 | **0.3%** | 53 | **0.4%** | 13 | **0.2%** |
|  | Trimester 2 | 1 | 1 | 81 | **0.4%** | 4 | **0.1%** | 58 | **0.4%** | 21 | **0.4%** |
|  | Trimester 2 | 2 | 0 | 617 | **2.7%** | 31 | **1.1%** | 499 | **3.6%** | 81 | **1.4%** |
|  | Trimester 2 | 2 | 1 | 108 | **0.5%** | 0 | **0.0%** | 538 | **3.9%** | 219 | **3.9%** |
|  | Trimester 3 | 1 | 0 | 525 | **2.3%** | 42 | **1.4%** | 123 | **0.9%** | 29 | **0.5%** |
|  | Trimester 3 | 1 | 1 | 1,272 | **5.5%** | 92 | **3.1%** | 713 | **5.1%** | 234 | **4.1%** |
|  | Trimester 3 | 1 | 2 | 42 | **0.2%** | 0 | **0.0%** | 398 | **2.9%** | 179 | **3.2%** |
|  | Trimester 3 | 2 | 0 | 1,750 | **7.6%** | 52 | **1.8%** | 878 | **6.3%** | 169 | **3.0%** |
|  | Trimester 3 | 2 | 1 | 113 | **0.5%** | 5 | **0.2%** | 640 | **4.6%** | 309 | **5.5%** |
|  | Trimester 3 | 3 | 0 | 55 | **0.2%** | 0 | **0.0%** | 140 | **1.0%** | 17 | **0.3%** |
| **Risk Status of mother** | None |  |  | 17,879 | **77.6%** | 2,276 | **77.8%** | 10,818 | **77.8%** | 4,428 | **78.1%** |
|  | At risk/CEV/Severely immunosuppressed | | | 5,174 | **22.4%** | 648 | **22.2%** | 3,090 | **22.2%** | 1,241 | **21.9%** |
| **Variant of most recent previous infection pre-birth in mother** | None |  |  | 20,236 | **87.8%** | 2,833 | **96.9%** | 11,951 | **85.9%** | 5,064 | **89.3%** |
|  | Wild-type |  |  | 753 | **3.3%** | 39 | **1.3%** | 470 | **3.4%** | 169 | **3.0%** |
|  | Alpha |  |  | 1,457 | **6.3%** | 44 | **1.5%** | 812 | **5.8%** | 254 | **4.5%** |
|  | Delta |  |  | 607 | **2.6%** | 8 | **0.3%** | 672 | **4.8%** | 182 | **3.2%** |
|  | Omicron |  |  | 0 | **0.0%** | 0 | **0.0%** | 3 | **0.0%** | 0 | **0.0%** |
| **Infection in the mother prior to infant infection** | None |  |  | 21,870 | **94.9%** | 1,999 | **68.4%** | 12,562 | **90.3%** | 4,610 | **81.3%** |
|  | More than 4 weeks prior |  |  | 678 | **2.9%** | 22 | **0.8%** | 953 | **6.9%** | 175 | **3.1%** |
|  | Less than 4 weeks prior |  |  | 505 | **2.2%** | 903 | **30.9%** | 393 | **2.8%** | 884 | **15.6%** |

Supplementary Table 9. Maternal vaccine effectiveness against symptomatic disease with Delta and Omicron in infants in England.

| Infant age (months) | Maternal Vaccination Status | Doses in pregnancy | Doses post pregnancy | Delta | | | Omicron | | |
| --- | --- | --- | --- | --- | --- | --- | --- | --- | --- |
|  |  |  |  | Cases | Controls | VE (95% CI) | Cases | Controls | VE (95% CI) |
| 0-5 | None | 0 | 0 | 1,609 | 10,065 | Baseline | 1,383 | 3,058 | Baseline |
|  | None | 0 | 1 | 554 | 3,633 | 34.5 (26.2 to 41.9) | 506 | 1,047 | 4.2 (-10.8 to 17.2) |
|  | None | 0 | 2 | 343 | 2,569 | 51.2 (42.6 to 58.4) | 604 | 1,446 | 25.8 (13.6 to 36.2) |
|  | Pre-pregnancy | 1 | 0 | 3 | 62 |  | 39 | 93 | -4.4 (-58.6 to 31.3) |
|  | Pre-pregnancy | 1 | 1 | 0 | 12 |  | 21 | 48 | 30.1 (-20.9 to 59.6) |
|  | Pre-pregnancy | 2 | 0 | 0 | 10 |  | 6 | 21 |  |
|  | Trimester 1 | 1 | 0 | 15 | 144 | 9.0 (-61.9 to 48.9) | 48 | 121 | -5.0 (-53.8 to 28.3) |
|  | Trimester 1 | 1 | 1 | 2 | 37 |  | 39 | 83 | 37.6 (5.7 to 58.6) |
|  | Trimester 1 | 2 | 0 | 3 | 70 | 65.3 (-15.7 to 89.6) | 40 | 105 | 3.9 (-44.4 to 36.0) |
|  | Trimester 1 | 2 | 1 | 2 | 19 |  | 37 | 98 | 44.0 (15.1 to 63.1) |
|  | Trimester 2 | 1 | 0 | 9 | 68 | -8.0 (-128.8 to 49.0) | 13 | 52 | 42.5 (-11.2 to 70.3) |
|  | Trimester 2 | 1 | 1 | 3 | 80 | 74.7 (16.6 to 92.3) | 18 | 53 | 29.6 (-28.3 to 61.4) |
|  | Trimester 2 | 2 | 0 | 31 | 617 | 66.7 (50.9 to 77.4) | 81 | 499 | 56.8 (43.4 to 67.0) |
|  | Trimester 2 | 2 | 1 | 0 | 105 |  | 203 | 511 | 58.0 (48.6 to 65.7) |
|  | Trimester 3 | 1 | 0 | 42 | 525 | 51.5 (31.9 to 65.4) | 28 | 119 | 35.8 (-2.0 to 59.6) |
|  | Trimester 3 | 1 | 1 | 90 | 1,203 | 70.7 (62.8 to 76.9) | 193 | 611 | 30.5 (14.6 to 43.5) |
|  | Trimester 3 | 1 | 2 | 0 | 23 |  | 58 | 108 | 52.9 (32.5 to 67.1) |
|  | Trimester 3 | 2 | 0 | 51 | 1,743 | 86.5 (81.9 to 90.0) | 163 | 867 | 56.6 (46.7 to 64.6) |
|  | Trimester 3 | 2 | 1 | 4 | 82 | 65.4 (0.6 to 87.9) | 224 | 481 | 56.2 (46.4 to 64.2) |
|  | Trimester 3 | 3 | 0 | 0 | 55 |  | 17 | 140 | 84.0 (72.7 to 90.6) |
| 0-2 | None | 0 | 0 | 1,220 | 7,858 | Baseline | 708 | 1,693 | Baseline |
|  | None | 0 | 1 | 357 | 2,408 | 37.7 (27.7 to 46.4) | 183 | 402 | 13.6 (-9.1 to 31.6) |
|  | None | 0 | 2 | 19 | 140 | 53.9 (22.1 to 72.7) | 17 | 29 | 25.9 (-52.4 to 64.0) |
|  | Pre-pregnancy | 1 | 0 | 3 | 61 |  | 34 | 84 | 1.8 (-56.2 to 38.2) |
|  | Pre-pregnancy | 1 | 1 | 0 | 12 |  | 14 | 35 | 41.1 (-15.2 to 69.9) |
|  | Pre-pregnancy | 2 | 0 | 0 | 10 |  | 5 | 18 |  |
|  | Trimester 1 | 1 | 0 | 15 | 132 | 4.1 (-72.3 to 46.6) | 40 | 100 | -12.2 (-73.1 to 27.2) |
|  | Trimester 1 | 1 | 1 | 0 | 32 |  | 18 | 44 | 56.4 (20.6 to 76.0) |
|  | Trimester 1 | 2 | 0 | 3 | 69 | 63.7 (-22.4 to 89.2) | 35 | 96 | 9.3 (-41.6 to 41.9) |
|  | Trimester 1 | 2 | 1 | 2 | 19 |  | 25 | 68 | 47.6 (12.4 to 68.6) |
|  | Trimester 2 | 1 | 0 | 8 | 58 | -18.3 (-169.3 to 48.0) | 10 | 37 | 26.8 (-59.3 to 66.4) |
|  | Trimester 2 | 1 | 1 | 1 | 47 | 83.4 (-27.1 to 97.8) | 8 | 22 |  |
|  | Trimester 2 | 2 | 0 | 27 | 589 | 70.1 (54.5 to 80.4) | 65 | 448 | 63.0 (49.5 to 72.9) |
|  | Trimester 2 | 2 | 1 | 0 | 67 |  | 76 | 288 | 72.3 (62.3 to 79.6) |
|  | Trimester 3 | 1 | 0 | 42 | 511 | 51.5 (31.7 to 65.5) | 18 | 96 | 46.7 (5.8 to 69.8) |
|  | Trimester 3 | 1 | 1 | 32 | 593 | 75.4 (64.0 to 83.2) | 43 | 149 | 43.6 (15.7 to 62.3) |
|  | Trimester 3 | 1 | 2 | 0 | 0 |  | 0 | 0 |  |
|  | Trimester 3 | 2 | 0 | 34 | 1,430 | 89.0 (84.3 to 92.4) | 89 | 573 | 63.5 (51.8 to 72.4) |
|  | Trimester 3 | 2 | 1 | 0 | 20 |  | 24 | 81 | 74.9 (58.4 to 84.8) |
|  | Trimester 3 | 3 | 0 | 0 | 55 |  | 12 | 122 | 87.4 (76.3 to 93.3) |
| 3-5 | None | 0 | 0 | 389 | 2,207 | Baseline | 675 | 1,365 | Baseline |
|  | None | 0 | 1 | 197 | 1,225 | 26.5 (9.9 to 40.1) | 323 | 645 | -3.7 (-25.2 to 14.1) |
|  | None | 0 | 2 | 324 | 2,429 | 48.4 (37.7 to 57.2) | 587 | 1,417 | 17.6 (2.8 to 30.2) |
|  | Pre-pregnancy | 1 | 0 | 0 | 1 |  | 5 | 9 |  |
|  | Pre-pregnancy | 1 | 1 | 0 | 0 |  | 7 | 13 |  |
|  | Pre-pregnancy | 2 | 0 | 0 | 0 |  | 1 | 3 |  |
|  | Trimester 1 | 1 | 0 | 0 | 12 |  | 8 | 21 |  |
|  | Trimester 1 | 1 | 1 | 2 | 5 |  | 21 | 39 | 16.4 (-49.0 to 53.1) |
|  | Trimester 1 | 2 | 0 | 0 | 1 |  | 5 | 9 |  |
|  | Trimester 1 | 2 | 1 | 0 | 0 |  | 12 | 30 | 45.4 (-13.2 to 73.7) |
|  | Trimester 2 | 1 | 0 | 1 | 10 |  | 3 | 15 | 68.0 (-17.1 to 91.3) |
|  | Trimester 2 | 1 | 1 | 2 | 33 |  | 10 | 31 |  |
|  | Trimester 2 | 2 | 0 | 4 | 28 |  | 16 | 51 | 34.8 (-21.4 to 64.9) |
|  | Trimester 2 | 2 | 1 | 0 | 38 |  | 127 | 223 | 42.7 (24.3 to 56.6) |
|  | Trimester 3 | 1 | 0 | 0 | 14 |  | 10 | 23 | 7.7 (-114.3 to 60.3) |
|  | Trimester 3 | 1 | 1 | 58 | 610 | 65.5 (52.6 to 74.9) | 150 | 462 | 20.9 (-1.1 to 38.1) |
|  | Trimester 3 | 1 | 2 | 0 | 23 |  | 58 | 108 | 44.1 (19.3 to 61.3) |
|  | Trimester 3 | 2 | 0 | 17 | 313 | 77.6 (62.0 to 86.8) | 74 | 294 | 47.7 (28.7 to 61.6) |
|  | Trimester 3 | 2 | 1 | 4 | 62 | 60.1 (-16.9 to 86.4) | 200 | 400 | 44.9 (30.7 to 56.3) |
|  | Trimester 3 | 3 | 0 | 0 | 0 |  | 5 | 18 | 75.5 (30.2 to 91.4) |
| 6-8 | None | 0 | 0 | - | - | - | 397 | 840 | Baseline |
|  | None | 0 | 1 | - | - | - | 146 | 320 | 6.6 (-20.6 to 27.7) |
|  | None | 0 | 2 | - | - | - | 1,132 | 2,582 | 9.9 (-6.0 to 23.5) |
|  | Pre-pregnancy | 1 | 0 | - | - | - | 0 | 2 |  |
|  | Pre-pregnancy | 1 | 1 | - | - | - | 0 | 0 |  |
|  | Pre-pregnancy | 2 | 0 | - | - | - | 0 | 0 |  |
|  | Trimester 1 | 1 | 0 | - | - | - | 0 | 1 |  |
|  | Trimester 1 | 1 | 1 | - | - | - | 0 | 2 |  |
|  | Trimester 1 | 2 | 0 | - | - | - | 0 | 0 |  |
|  | Trimester 1 | 2 | 1 | - | - | - | 0 | 1 |  |
|  | Trimester 2 | 1 | 0 | - | - | - | 0 | 1 |  |
|  | Trimester 2 | 1 | 1 | - | - | - | 3 | 5 |  |
|  | Trimester 2 | 2 | 0 | - | - | - | 0 | 0 |  |
|  | Trimester 2 | 2 | 1 | - | - | - | 16 | 27 | 22.5 (-52.7 to 60.7) |
|  | Trimester 3 | 1 | 0 | - | - | - | 1 | 4 |  |
|  | Trimester 3 | 1 | 1 | - | - | - | 41 | 102 | 19.3 (-24.2 to 47.6) |
|  | Trimester 3 | 1 | 2 | - | - | - | 121 | 290 | 45.1 (27.7 to 58.3) |
|  | Trimester 3 | 2 | 0 | - | - | - | 6 | 11 |  |
|  | Trimester 3 | 2 | 1 | - | - | - | 85 | 159 | 30.1 (2.8 to 49.7) |
|  | Trimester 3 | 3 | 0 | - | - | - | 0 | 0 |  |

Supplementary Table 10. Protection of maternal past infection in the mother against symptomatic disease with Delta and Omicron in infants in England.

| Infant age (months) | Maternal Previous Infection | Delta | | | | Omicron | | |
| --- | --- | --- | --- | --- | --- | --- | --- | --- |
|  |  | Cases | Controls | Protection against Delta (95% CI) | Cases | | Controls | Protection against Omicron (95% CI) |
| 0-5 | No | 2684 | 18483 | Baseline | 3269 | | 8010 | Baseline |
|  | Wild-type | 34 | 699 | 70.3 (57.4 to 79.2) | 118 | | 339 | 23.7 (3.1 to 39.9) |
|  | Alpha | 35 | 1333 | 83.3 (76.4 to 88.1) | 162 | | 569 | 36.5 (22.4 to 47.9) |
|  | Delta | 8 | 607 | 87.7 (75.0 to 93.9) | 172 | | 640 | 41.1 (28.7 to 51.3) |
| 0-2 | No | 1723 | 12208 | Baseline | 1234 | | 3532 | Baseline |
|  | Wild-type | 14 | 468 | 81.8 (68.5 to 89.5) | 45 | | 151 | 28.6 (-4.7 to 51.3) |
|  | Alpha | 21 | 903 | 85.1 (76.8 to 90.4) | 61 | | 263 | 38.6 (15.5 to 55.4) |
|  | Delta | 5 | 532 | 91.8 (80.1 to 96.7) | 84 | | 436 | 54.1 (40.1 to 64.9) |
| 3-5 | No | 1110 | 8028 | Baseline | 2035 | | 4478 | Baseline |
|  | Wild-type | 25 | 285 | 47.7 (14.6 to 68.0) | 73 | | 188 | 20.3 (-8.8 to 41.5) |
|  | Alpha | 23 | 554 | 80.3 (65.8 to 88.6) | 101 | | 306 | 34.8 (15.6 to 49.6) |
|  | Delta | 3 | 75 | 49.1 (-68.9 to 84.7) | 88 | | 204 | 23.0 (-1.8 to 41.8) |
| 6-8 | No | - | - | - | 1795 | | 3941 | Baseline |
|  | Wild-type | - | - | - | 51 | | 131 | 16.1 (-20.1 to 41.4) |
|  | Alpha | - | - | - | 92 | | 243 | 17.2 (-8.2 to 36.6) |
|  | Delta | - | - | - | 10 | | 32 | 49.1 (-9.6 to 76.3) |

Supplementary Table 11. Descriptive characteristics of hospitalised infants for whom vaccine effectiveness was estimated.

|  |  |  |  | **Delta period** | | | | **Omicron period** | | | |
| --- | --- | --- | --- | --- | --- | --- | --- | --- | --- | --- | --- |
|  | **Test Result** |  |  | **Control (Negative)** | | **Case (Positive)** | | **Control (Negative)** | | **Case (Positive)** | |
|  |  |  |  | **n** | **%** | **n** | **%** | **n** | **%** | **n** | **%** |
| **Pillar** | 1 |  |  | 4,588 | **91.3%** | 436 | **8.7%** | 1,413 | **75.6%** | 457 | **24.4%** |
| **Gender of infant** | Female |  |  | 2,009 | **43.8%** | 184 | **42.2%** | 529 | **37.4%** | 196 | **42.9%** |
|  | Male |  |  | 2,547 | **55.5%** | 252 | **57.8%** | 873 | **61.8%** | 261 | **57.1%** |
|  | Unknown |  |  | 32 | **0.7%** | 0 | **0.0%** | 11 | **0.8%** | 0 | **0.0%** |
| **Premature** | No |  |  | 3,963 | **86.4%** | 405 | **92.9%** | 1,192 | **84.4%** | 406 | **88.8%** |
|  | Gestational age <37 weeks | |  | 450 | **9.8%** | 26 | **6.0%** | 135 | **9.6%** | 33 | **7.2%** |
|  | Gestational age <33 weeks | |  | 175 | **3.8%** | 5 | **1.1%** | 86 | **6.1%** | 18 | **3.9%** |
| **Age of infant (months)** | 0 |  |  | 652 | **14.2%** | 148 | **33.9%** | 77 | **5.4%** | 45 | **9.8%** |
|  | 1 |  |  | 1,878 | **40.9%** | 188 | **43.1%** | 350 | **24.8%** | 138 | **30.2%** |
|  | 2 |  |  | 945 | **20.6%** | 74 | **17.0%** | 229 | **16.2%** | 111 | **24.3%** |
|  | 3 |  |  | 530 | **11.6%** | 13 | **3.0%** | 226 | **16.0%** | 64 | **14.0%** |
|  | 4 |  |  | 340 | **7.4%** | 12 | **2.8%** | 237 | **16.8%** | 46 | **10.1%** |
|  | 5 |  |  | 243 | **5.3%** | 1 | **0.2%** | 294 | **20.8%** | 53 | **11.6%** |
| **Ethnicity** | African |  |  | 86 | **1.9%** | 7 | **1.6%** | 28 | **2.0%** | 10 | **2.2%** |
|  | Any other Asian background | |  | 79 | **1.7%** | 11 | **2.5%** | 42 | **3.0%** | 13 | **2.8%** |
|  | Any other Black background | |  | 33 | **0.7%** | 3 | **0.7%** | 15 | **1.1%** | 3 | **0.7%** |
|  | Any other White background | |  | 243 | **5.3%** | 57 | **13.1%** | 71 | **5.0%** | 85 | **18.6%** |
|  | Any other ethnic group | |  | 84 | **1.8%** | 15 | **3.4%** | 31 | **2.2%** | 14 | **3.1%** |
|  | Any other mixed background | |  | 70 | **1.5%** | 6 | **1.4%** | 24 | **1.7%** | 10 | **2.2%** |
|  | Bangladeshi or British Bangladeshi | | | 67 | **1.5%** | 12 | **2.8%** | 18 | **1.3%** | 3 | **0.7%** |
|  | British, Mixed British | |  | 2,637 | **57.5%** | 183 | **42.0%** | 915 | **64.8%** | 219 | **47.9%** |
|  | Caribbean |  |  | 12 | **0.3%** | 6 | **1.4%** | 1 | **0.1%** | 1 | **0.2%** |
|  | Chinese |  |  | 7 | **0.2%** | 0 | **0.0%** | 2 | **0.1%** | 0 | **0.0%** |
|  | Indian or British Indian | |  | 77 | **1.7%** | 13 | **3.0%** | 28 | **2.0%** | 22 | **4.8%** |
|  | Irish |  |  | 14 | **0.3%** | 2 | **0.5%** | 6 | **0.4%** | 0 | **0.0%** |
|  | Pakistani or British Pakistani | | | 186 | **4.1%** | 20 | **4.6%** | 87 | **6.2%** | 27 | **5.9%** |
|  | White and Asian | |  | 33 | **0.7%** | 5 | **1.1%** | 11 | **0.8%** | 7 | **1.5%** |
|  | White and Black African | |  | 24 | **0.5%** | 4 | **0.9%** | 6 | **0.4%** | 2 | **0.4%** |
|  | White and Black Caribbean | |  | 43 | **0.9%** | 7 | **1.6%** | 19 | **1.3%** | 8 | **1.8%** |
|  | Missing |  |  | 893 | **19.5%** | 85 | **19.5%** | 109 | **7.7%** | 33 | **7.2%** |
| **IMD Quintiles** | 1 |  |  | 1,347 | **29.4%** | 136 | **31.2%** | 452 | **32.0%** | 123 | **26.9%** |
|  | 2 |  |  | 887 | **19.3%** | 84 | **19.3%** | 317 | **22.4%** | 109 | **23.9%** |
|  | 3 |  |  | 767 | **16.7%** | 91 | **20.9%** | 219 | **15.5%** | 96 | **21.0%** |
|  | 4 |  |  | 784 | **17.1%** | 77 | **17.7%** | 231 | **16.3%** | 82 | **17.9%** |
|  | 5 |  |  | 761 | **16.6%** | 44 | **10.1%** | 185 | **13.1%** | 45 | **9.8%** |
|  | Missing |  |  | 42 | **0.9%** | 4 | **0.9%** | 9 | **0.6%** | 2 | **0.4%** |
| **Age of mother (years)** | 15-24 |  |  | 724 | **15.8%** | 78 | **17.9%** | 210 | **14.9%** | 95 | **20.8%** |
|  | 25-29 |  |  | 1,358 | **29.6%** | 126 | **28.9%** | 420 | **29.7%** | 125 | **27.4%** |
|  | 30-34 |  |  | 1,514 | **33.0%** | 136 | **31.2%** | 471 | **33.3%** | 139 | **30.4%** |
|  | 35+ |  |  | 992 | **21.6%** | 96 | **22.0%** | 312 | **22.1%** | 98 | **21.4%** |
| **Mother's vaccination status; no. of doses in pregnancy and no. of doses post pregnancy** | Unvaccinated | 0 | 0 | 2,489 | **54.3%** | 341 | **78.2%** | 560 | **39.6%** | 270 | **59.1%** |
|  | Unvaccinated | 0 | 1 | 926 | **20.2%** | 64 | **14.7%** | 159 | **11.3%** | 59 | **12.9%** |
|  | Unvaccinated | 0 | 2 | 318 | **6.9%** | 7 | **1.6%** | 147 | **10.4%** | 19 | **4.2%** |
|  | Trimester 1 | 1 | 0 | 34 | **0.7%** | 4 | **0.9%** | 26 | **1.8%** | 6 | **1.3%** |
|  | Trimester 1 | 1 | 1 | 8 | **0.2%** | 0 | **0.0%** | 17 | **1.2%** | 3 | **0.7%** |
|  | Trimester 1 | 2 | 0 | 9 | **0.2%** | 0 | **0.0%** | 17 | **1.2%** | 10 | **2.2%** |
|  | Trimester 1 | 2 | 1 | 5 | **0.1%** | 0 | **0.0%** | 25 | **1.8%** | 3 | **0.7%** |
|  | Trimester 2 | 1 | 0 | 25 | **0.5%** | 1 | **0.2%** | 21 | **1.5%** | 2 | **0.4%** |
|  | Trimester 2 | 1 | 1 | 25 | **0.5%** | 0 | **0.0%** | 10 | **0.7%** | 1 | **0.2%** |
|  | Trimester 2 | 2 | 0 | 108 | **2.4%** | 2 | **0.5%** | 87 | **6.2%** | 13 | **2.8%** |
|  | Trimester 2 | 2 | 1 | 14 | **0.3%** | 0 | **0.0%** | 76 | **5.4%** | 24 | **5.3%** |
|  | Trimester 3 | 1 | 0 | 149 | **3.2%** | 10 | **2.3%** | 28 | **2.0%** | 3 | **0.7%** |
|  | Trimester 3 | 1 | 1 | 187 | **4.1%** | 5 | **1.1%** | 54 | **3.8%** | 12 | **2.6%** |
|  | Trimester 3 | 1 | 2 | 1 | **0.0%** | 0 | **0.0%** | 8 | **0.6%** | 3 | **0.7%** |
|  | Trimester 3 | 2 | 0 | 273 | **6.0%** | 2 | **0.5%** | 114 | **8.1%** | 14 | **3.1%** |
|  | Trimester 3 | 2 | 1 | 9 | **0.2%** | 0 | **0.0%** | 41 | **2.9%** | 12 | **2.6%** |
|  | Trimester 3 | 3 | 0 | 8 | **0.2%** | 0 | **0.0%** | 23 | **1.6%** | 3 | **0.7%** |
| **Risk Status of mother** | None |  |  | 3,453 | **75.3%** | 352 | **80.7%** | 1,075 | **76.1%** | 369 | **80.7%** |
|  | At risk/CEV/Severely immunosuppressed | | | 1,135 | **24.7%** | 84 | **19.3%** | 338 | **23.9%** | 88 | **19.3%** |
| **Variant of most recent previous infection pre-birth in mother** | None |  |  | 4,021 | **87.6%** | 420 | **96.3%** | 1,173 | **83.0%** | 390 | **85.3%** |
|  | Wild-type |  |  | 141 | **3.1%** | 1 | **0.2%** | 52 | **3.7%** | 15 | **3.3%** |
|  | Alpha |  |  | 299 | **6.5%** | 13 | **3.0%** | 87 | **6.2%** | 24 | **5.3%** |
|  | Delta |  |  | 127 | **2.8%** | 2 | **0.5%** | 100 | **7.1%** | 28 | **6.1%** |
|  | Omicron |  |  | 0 | **0.0%** | 0 | **0.0%** | 1 | **0.1%** | 0 | **0.0%** |

Supplementary Table 12. Maternal vaccine effectiveness against hospitalisation with Delta and Omicron in infants in England.

| Infant age (months) | Maternal Vaccination Status | Doses in pregnancy | Doses post pregnancy | Delta | | | Omicron | | |
| --- | --- | --- | --- | --- | --- | --- | --- | --- | --- |
|  |  |  |  | Cases | Controls | VE (95% CI) | Cases | Controls | VE (95% CI) |
| 0-6 | None | 0 | 0 | 341 | 2,489 | Baseline | 270 | 560 | Baseline |
|  | None | 0 | 1 | 64 | 926 | 34.0 (9.9 to 51.7) | 59 | 159 | 6.5 (-45.1 to 39.7) |
|  | None | 0 | 2 | 7 | 318 | 26.3 (-74.5 to 68.8) | 19 | 147 | 52.0 (10.2 to 74.3) |
|  | Trimester 1 | 1 | 0 | 4 | 34 |  | 6 | 26 |  |
|  | Trimester 1 | 1 | 1 | 0 | 8 |  | 3 | 17 |  |
|  | Trimester 1 | 2 | 0 | 0 | 9 |  | 10 | 17 |  |
|  | Trimester 1 | 2 | 1 | 0 | 5 |  | 3 | 25 | 86.4 (47.0 to 96.5) |
|  | Trimester 2 | 1 | 0 | 1 | 25 |  | 2 | 21 | 84.6 (17.8 to 97.1) |
|  | Trimester 2 | 1 | 1 | 0 | 25 |  | 1 | 10 |  |
|  | Trimester 2 | 2 | 0 | 2 | 108 | 87.4 (47.2 to 97.0) | 13 | 87 | 73.9 (43.9 to 87.9) |
|  | Trimester 2 | 2 | 1 | 0 | 14 |  | 24 | 76 | 67.8 (41.5 to 82.3) |
|  | Trimester 3 | 1 | 0 | 10 | 149 | 54.8 (10.6 to 77.1) | 3 | 28 | 84.1 (30.5 to 96.4) |
|  | Trimester 3 | 1 | 1 | 5 | 187 | 76.5 (40.7 to 90.7) | 12 | 54 | 55.5 (5.2 to 79.2) |
|  | Trimester 3 | 1 | 2 | 0 | 1 |  | 3 | 8 |  |
|  | Trimester 3 | 2 | 0 | 2 | 273 | 94.7 (78.2 to 98.7) | 14 | 114 | 78.7 (58.2 to 89.1) |
|  | Trimester 3 | 2 | 1 | 0 | 9 |  | 12 | 41 | 56.0 (6.3 to 79.4) |
|  | Trimester 3 | 3 | 0 | 0 | 8 |  | 3 | 23 | 90.5 (62.4 to 97.6) |

Supplementary Table 13. Protection of maternal past infection in the mother against hospitalisation with Delta and Omicron in infants in England.

| Infant age (months) | Maternal Previous Infection | Delta | | | Omicron | | |
| --- | --- | --- | --- | --- | --- | --- | --- |
|  |  | Cases | Controls | Protection against Delta (95% CI) | Cases | Controls | Protection against Omicron (95% CI) |
| 0-6 | No | 420 | 4021 | Baseline | 390 | 1173 | Baseline |
|  | Wild-type | 1 | 141 | 93.4 (52.0 to 99.1) | 15 | 52 | 8.4 (-85.5 to 54.8) |
|  | Alpha | 13 | 299 | 63.3 (33.9 to 79.7) | 24 | 87 | 11.2 (-61.9 to 51.3) |
|  | Delta | 2 | 127 | 85.1 (38.0 to 96.4) | 28 | 100 | 24.0 (-30.1 to 55.6) |

3.	UK Health Security Agency. COVID-19: the green book, chapter 14a. Immunisation against infectious diseases: UK Health Security Agency,; 2020.
